# Micro-fragmented adipose tissue reprogrammes the systemic molecular landscape of inflammatory knee osteoarthritis

**DOI:** 10.64898/2026.06.12.26355510

**Authors:** Dragan Primorac, Vilim Molnar, Petar Brlek, Luka Bulić, Željko Jeleč, Uršula Prosenc Zmrzljak, Thomas Klarić, Gordan Lauc, Noël Malod-Dognin, Nataša Pržulj

**Affiliations:** St. Catherine Specialty Hospital, 10000 Zagreb, Croatia; School of Medicine, Josip Juraj Strossmayer University of Osijek, 31000 Osijek, Croatia; International Center for Applied Biological Research, 10000 Zagreb, Croatia; Faculty of Dental Medicine and Health, Josip Juraj Strossmayer University of Osijek, 31000 Osijek, Croatia; Eberly College of Science, The Pennsylvania State University, State College, PA 16802, USA; The Henry C. Lee College of Criminal Justice and Forensic Sciences, University of New Haven, New Haven, CT 06516, USA; Sana Kliniken Oberfranken, 96450 Coburg, Germany; School of Medicine, University of Split, 21000 Split, Croatia; School of Medicine, University of Rijeka, 51000 Rijeka, Croatia; School of Medicine, University of Pittsburgh, Pittsburgh, PA 15213, USA; Gandhinagar Campus, National Forensic Sciences University, Gandhinagar 382007, India; Department of Molecular Biology, Faculty of Science, University of Zagreb, 10000 Zagreb, Croatia; Algebra Bernays University, 10000 Zagreb, Croatia; Department of Physiotherapy, University North, 42000 Varaždin, Croatia; Molecular Biology Laboratory, BIA Separations CRO, Labena Ltd., 1000 Ljubljana, Slovenia; Genos Glycoscience Research Laboratory, 10000 Zagreb, Croatia; Faculty of Pharmacy and Biochemistry, University of Zagreb, 10000 Zagreb, Croatia; Mohamed bin Zayed University of Artificial Intelligence, Masdar City, 00000 Abu Dhabi, United Arab Emirates; Department of Computer Science, University College London, London WC1E 6BT, United Kingdom

**Author notes:** Correspondence, Prof. Dragan Primorac, M.D., Ph.D., St. Catherine Specialty Hospital, Knez Branimir street 71E, 10000 Zagreb, Croatia.

**Keywords:** knee osteoarthritis, micro-fragmented adipose tissue, hyaluronic acid, multi-omics data fusion, non-negative matrix tri-factorization, molecular signatures, plasma biomarkers

## Abstract

Knee osteoarthritis affects an estimated 374 million people and has no approved disease-modifying treatment. In our double-blind randomised trial (ISRCTN88966184), intra-articular micro-fragmented adipose tissue (MFAT) outperformed hyaluronic acid (HA) on pain, function and cartilage composition — but clinical endpoints alone cannot say whether the disease changed or only the symptoms. Here we answer that question from the blood. Using an interpretable AI data-fusion framework based on non-negative matrix tri-factorisation, we integrated longitudinal plasma proteomics, N-glycomics and miRNA transcriptomics with patient genetics and prior molecular networks at baseline, one and six months, resolving 93 gene and 30 miRNA pathways, almost all significantly enriched in Gene Ontology Biological Process and Reactome annotations. By six months the two injections had left sharply distinct molecular fingerprints. HA remained dominated by canonical OA pathogenesis, including the cartilage-degrading effectors MMP13 and LIMK2 and persistent markers of synovial inflammation. MFAT instead shifted the systemic landscape towards chondroprotection, anti-inflammatory signalling and bone–cartilage homeostasis, prioritising effectors including SIRT7 and NDUFC1. These are, to our knowledge, the first in vivo human data to explain why a regenerative therapy outperforms viscosupplementation in knee osteoarthritis, and they place MFAT on the disease-modifying rather than the symptomatic side of that divide.

## 1. INTRODUCTION

Knee osteoarthritis (KOA) affects an estimated 374 million people worldwide and remains without an approved disease-modifying drug; clinical care is divided between symptomatic agents and arthroplasty ^1,2^. KOA is now recognised not as a single entity but as a constellation of biologically distinct phenotypes, of which the inflammatory phenotype, defined by synovitis and joint effusion in the absence of mechanical malalignment, is a particularly tractable target for biological intervention because synovial inflammation is a proximal driver of both pain and structural progression ^3,4^.

Intra-articular autologous micro-fragmented adipose tissue (MFAT), which delivers mesenchymal stromal cells within their native stromal vascular niche, has shown clinical benefit in inflammatory-phenotype KOA ^5–7^. In our recently completed double-blind randomized trial, MFAT outperformed hyaluronic acid (HA) on patient-reported symptom scores regarding effusion, stiffness and mobility ^8^. The molecular basis of this differential efficacy, however, remains unknown. Mechanistic insight is largely inferred from in vitro paracrine assays, and conventional plasma biomarker panels, such as single cytokines or isolated miRNAs, capture only narrow facets of a multi-systemic disease.

Notably, HA also produces clinically meaningful symptomatic and structural improvements in our trial and in the broader literature, supporting its continued role as a standard-of-care comparator, although guideline recommendations remain heterogeneous and effect sizes vary across patient subgroups ^9^. Preclinical work has attributed HA action to viscosupplementation, chondroprotection and modulation of inflammatory cytokines, while MFAT is thought to act through paracrine immunomodulation by resident MSCs ^10^. The two interventions thus appear to deliver overlapping but unequal clinical benefits through entirely different putative mechanisms. However, to our knowledge, no study has directly compared their in vivo molecular response in human osteoarthritis. This gap is consequential: distinguishing a treatment that modifies disease biology from one that merely relieves symptoms is the central unmet question in KOA, and without an in vivo molecular comparison neither rational patient selection nor a regulatory case for disease modification can be built.

Over the past decade, artificial intelligence (AI) has become central to biomedical research, driven by the accumulation of high-throughput molecular data and the recognition that complex diseases are rarely reducible to a single gene or pathway. The field has shifted from analyzing individual molecular layers in isolation toward integrative, systems-level approaches that jointly model genomic, transcriptomic, proteomic, glycomic, and metabolomic data alongside clinical phenotypes. The integration of multi-omics data with detailed phenotypic insights marks a paradigm shift in biomedical research. AI methods are well-suited to this setting because they handle high-dimensional, heterogeneous, and partially missing data while uncovering patterns that conventional pipelines miss, and they now span classical supervised learning, deep representation learning, graph neural networks, and matrix factorization techniques for multi-omics fusion ^11^.

Among these approaches, non-negative matrix tri-factorization (NMTF) is particularly well suited to the constraints outlined above. NMTF jointly decomposes multiple data matrices into shared low-dimensional factors, allowing heterogeneous omics layers and prior network knowledge to be fused within a single model rather than concatenated, so that the relational structure of each layer is preserved rather than discarded ^12^. Because it learns by clustering entities into interpretable groups and recovers latent associations directly from the shared factors, it yields biologically transparent modules instead of black-box predictions, and its reliance on collective patterns across data sources rather than on large numbers of training examples makes it robust at the modest cohort sizes typical of translational studies. These properties have made NMTF an effective tool for integrating and mining multi-omics data across various fields of biomedical research ^13–15^, motivating its application here to the comparison of MFAT and HA in inflammatory KOA.

In this study, we apply an interpretable AI-based multi-omics data-fusion framework to longitudinally collected plasma samples from this randomized cohort (ISRCTN88966184), integrating proteomics, N-glycomics, miRNA transcriptomics and whole-genome sequencing data with public protein–protein and miRNA–gene regulatory networks at baseline, one and six months ^16,17^. We identify functional modules of genes and miRNAs that diverge significantly between treatment arms by six months: HA leaves a persistent molecular signature dominated by canonical OA pathogenic pathways, including cartilage-degrading effectors and synovial inflammation, whereas MFAT shifts the systemic landscape toward chondroprotection, anti-inflammatory signalling and bone–cartilage homeostasis. These results provide the first systems-level molecular evidence that MFAT acts as a disease-modifying intervention in inflammatory KOA.

## 2. RESULTS

In the randomized trial from which this cohort is drawn, MFAT produced superior improvements in pain, function and cartilage composition (dGEMRIC) relative to HA ^8^. Why two intra-articular treatments with such different clinical trajectories diverge at the level of disease biology has, however, remained unknown, because their *in vivo* molecular responses have never been measured or compared in human osteoarthritis. We therefore set out to provide that missing molecular layer: using longitudinal plasma collected from the same trial participants, we compared the systems-level molecular response to MFAT and HA across baseline, one and six months, and asked whether the two treatments engage distinct biological programs. The analyses below establish, to our knowledge, for the first time in humans, the molecular correlates that distinguish a regenerative from a viscosupplementary intervention in KOA.

### 2.1. Cohort & sampling design

Between February 2020 and May 2023, 18,589 patients presenting to orthopedic outpatient clinics at three participating centres (St. Catherine Specialty Hospital, Zagreb; University Hospital Centre Sestre Milosrdnice, Zagreb; University Hospital Merkur, Zagreb) were screened for symptomatic knee osteoarthritis. Of these, 339 patients underwent a detailed eligibility assessment against the inflammatory-phenotype criteria defined below (Section 5.1), and 53 patients met all inclusion and exclusion criteria and were enrolled. Participants were randomized 2:1 to receive a single intra-articular injection of autologous micro-fragmented adipose tissue (MFAT; n = 35) or hyaluronic acid (HA; n = 18) (Figure 1A). Baseline demographic characteristics were comparable between arms: mean age 53.9 ± 9.0 years in the MFAT arm and 58.7 ± 9.5 years in the HA arm (p = 0.104); proportion of women 74.3% versus 77.8% (p = 0.780); body mass index 26.6 ± 2.4 versus 26.8 ± 2.5 kg/m² (p = 0.851).

**Figure 1.**
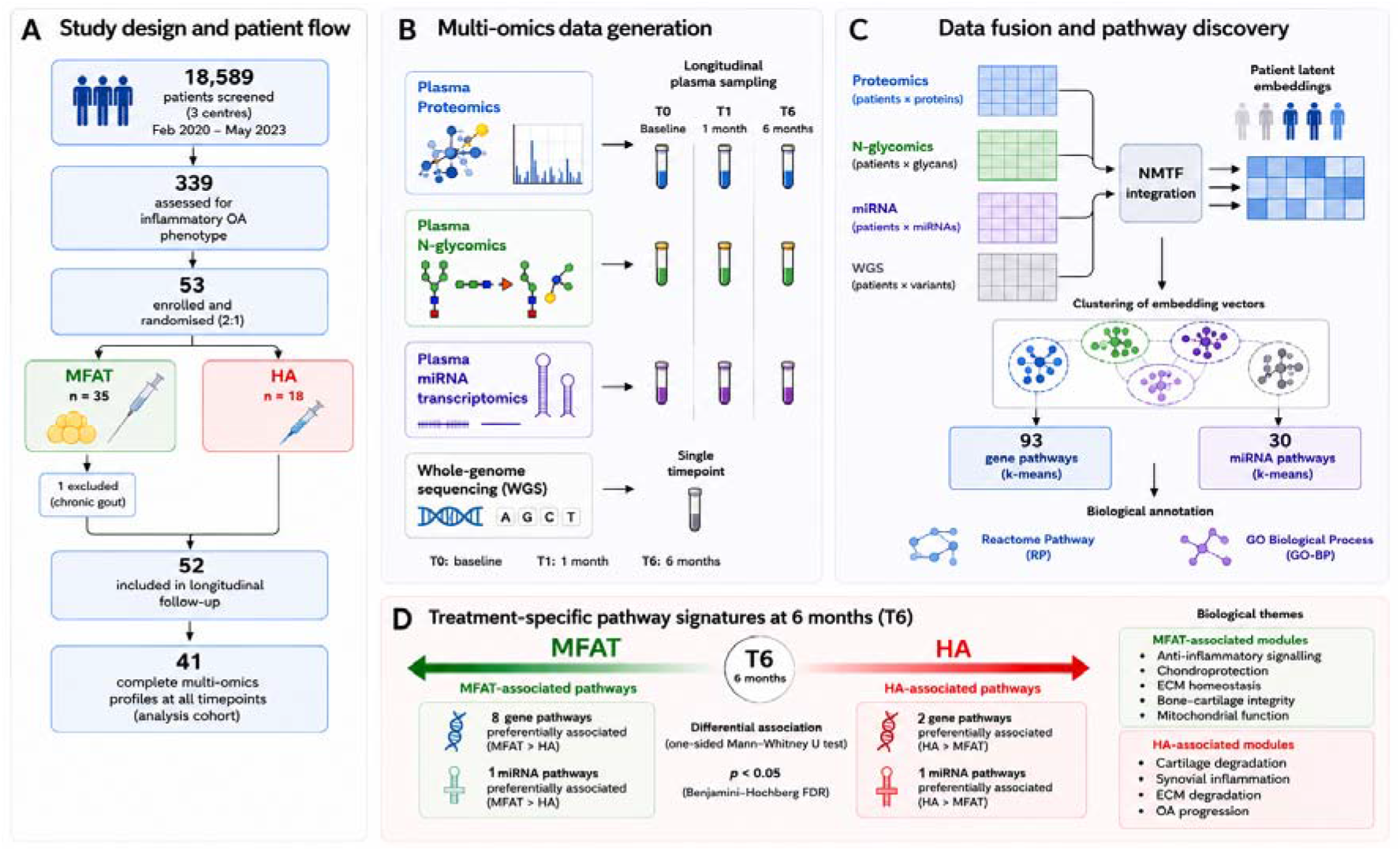
Study design, multi-omics integration, data fusion, and identification of treatment-associated molecular signatures. **(A)** Patient recruitment and cohort derivation between February 2020 and May 2023, including screening 18,589 orthopedic patients. **(B)** Multi-omics data generation. Plasma proteomics, plasma N-glycomics, and plasma miRNA transcriptomics were profiled longitudinally at baseline (T0), one month (T1), and six months (T6) following treatment. Germline whole-genome sequencing (WGS) was performed once per participant. **(C)** NMTF-based multi-omics data fusion and pathway discovery workflow, which identified 93 data-driven gene pathways and 30 data-driven miRNA pathways. **(D)** Identification of treatment-associated molecular signatures, resulting in ten gene pathways and two miRNA pathways exhibiting significant treatment-specific associations.

One participant from the MFAT arm was excluded from all post-intervention analyses following a confirmed diagnosis of chronic gout identified after randomisation. Multi-omics profiling comprised plasma proteomics, plasma N-glycomics, plasma miRNA transcriptomics, and whole-genome sequencing (Figure 1B). Plasma-derived molecular profiles were generated at baseline (T0), one month (T1), and six months (T6) following intervention, whereas whole-genome sequencing was conducted once per participant. Complete multi-omics profiles across all timepoints were available for 41 of the 53 enrolled patients, comprising the analysis cohort for data fusion.

### 2.2. Data-driven pathway discovery and functional assessment

Using gene and miRNA embedding vectors derived from our NMTF-based data fusion methodology, we identified 93 data-driven gene pathways and 30 miRNA pathways via k-means clustering of concatenated, timepoint-specific embedding vectors across all three timepoints (Figure 1C). To evaluate the biological relevance of these pathways, we assessed their enrichment in Reactome Pathway annotations.

As presented in Figure 2 (panels A and B), our data-driven pathways are almost all statistically significantly enriched in biological processes and pathways, with 96.7% of the data-driven pathways of genes and 86.7% of the data-driven pathways of miRNAs being statistically significantly enriched in at least one Reactome Pathway (RP) annotation. Furthermore, our data-driven pathways also cover well the biological function space, with 79.3% of the RP annotations being enriched in at least one pathway of genes and 93.6% of them being enriched in at least one pathway of miRNAs. Also, we observe similar enrichment results when using Gene Ontology Biological Process (GO-BP) annotations (see Figure 2, panels C and D). Importantly, as presented in Figure 2, such large enrichments are not observed in randomly generated pathways (clusters) having the same sizes.

**Figure 2.**
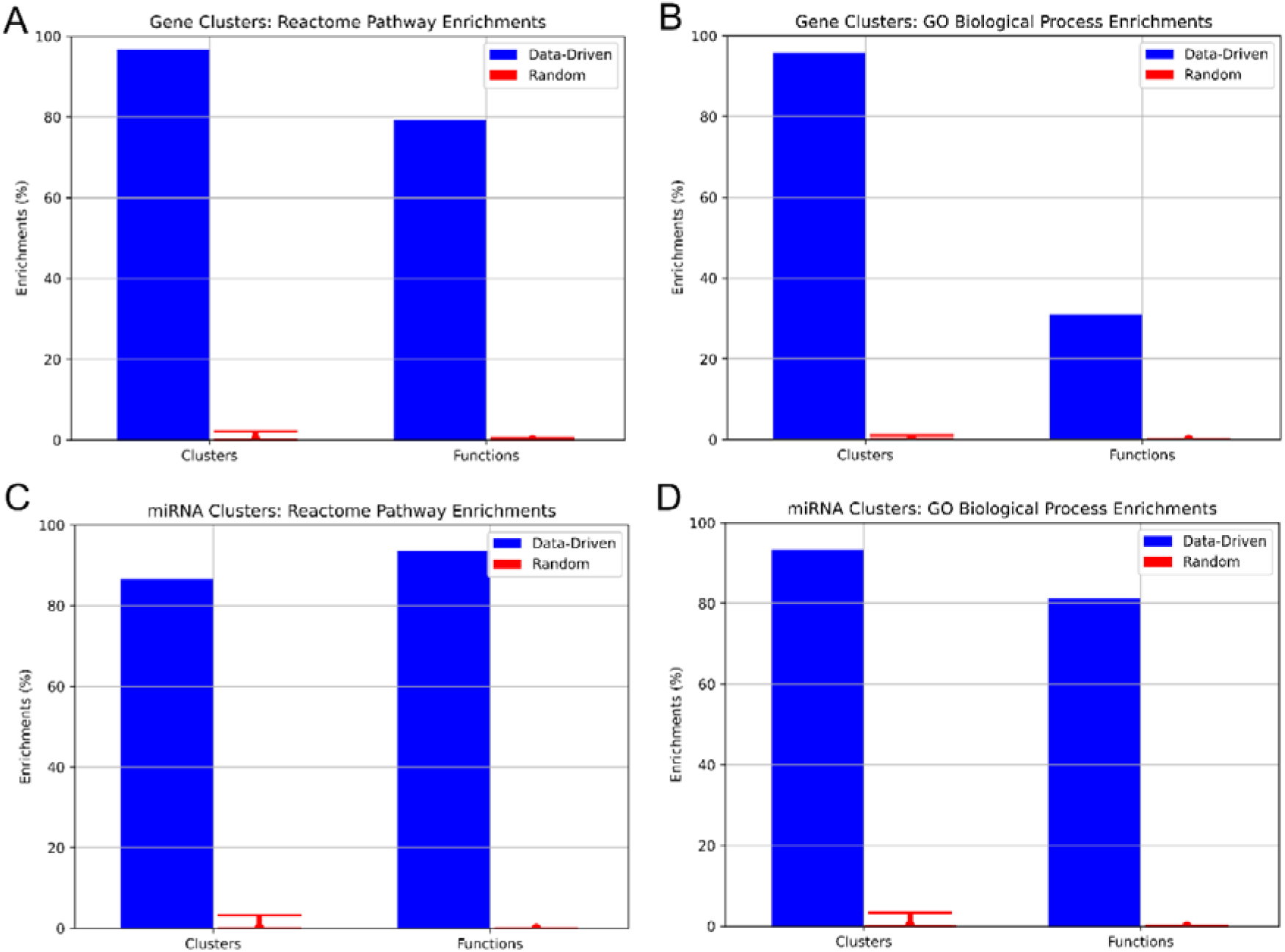
Functional enrichment analysis of the data-driven pathways. **Panel A:** For clusters (data-driven pathways) of genes, the blue bars show the percentage of the clusters having at least one Reactome Pathway (RP) annotation enriched in them (Clusters) and the percentage of the RP annotations that are enriched in at least one cluster (Functions). The red error bars show the corresponding minimum, average and maximum enrichment scores obtained over 25 random clusterings with the same cluster sizes. **Panels B, C and D** show the same, but for the enrichments of the gene clusters using Gene Ontology Biological Process (GO-BP) annotations, the enrichments of the miRNA clusters using RP annotations and the enrichments of the miRNA clusters using GO-BP annotations, respectively.

This confirms that our data-driven pathways capture biologically coherent functional modules, which emerged from the fusion of the data. In the next sections, we use these data-driven pathways to investigate the molecular basis of the MFAT and HA treatments.

### 2.3. Uncovering pathway-level signatures of MFAT and HA treatments

To identify molecular differences between MFAT and HA treatments, we leveraged the reconstructed patient–gene (*R_4_*) and patient–miRNA (*R_5_*) association matrices derived from our NMTF data fusion framework to determine which data-driven pathways were preferentially associated with each treatment group at each timepoint. Pathways showing statistically significant differential association were identified using a one-sided Mann–Whitney U test, with significance defined as p < 0.05 after Benjamini–Hochberg correction for multiple hypothesis testing (Figure 1D).

As presented in Table 1, ten out of our 93 data-driven pathways of genes (Table 1A) and two out of our 30 data-driven pathways of miRNAs (Table 1B) are significantly preferentially associated with the MFAT-treated patients or with the HA-treated patients at timepoint T6.

**Table 1.**
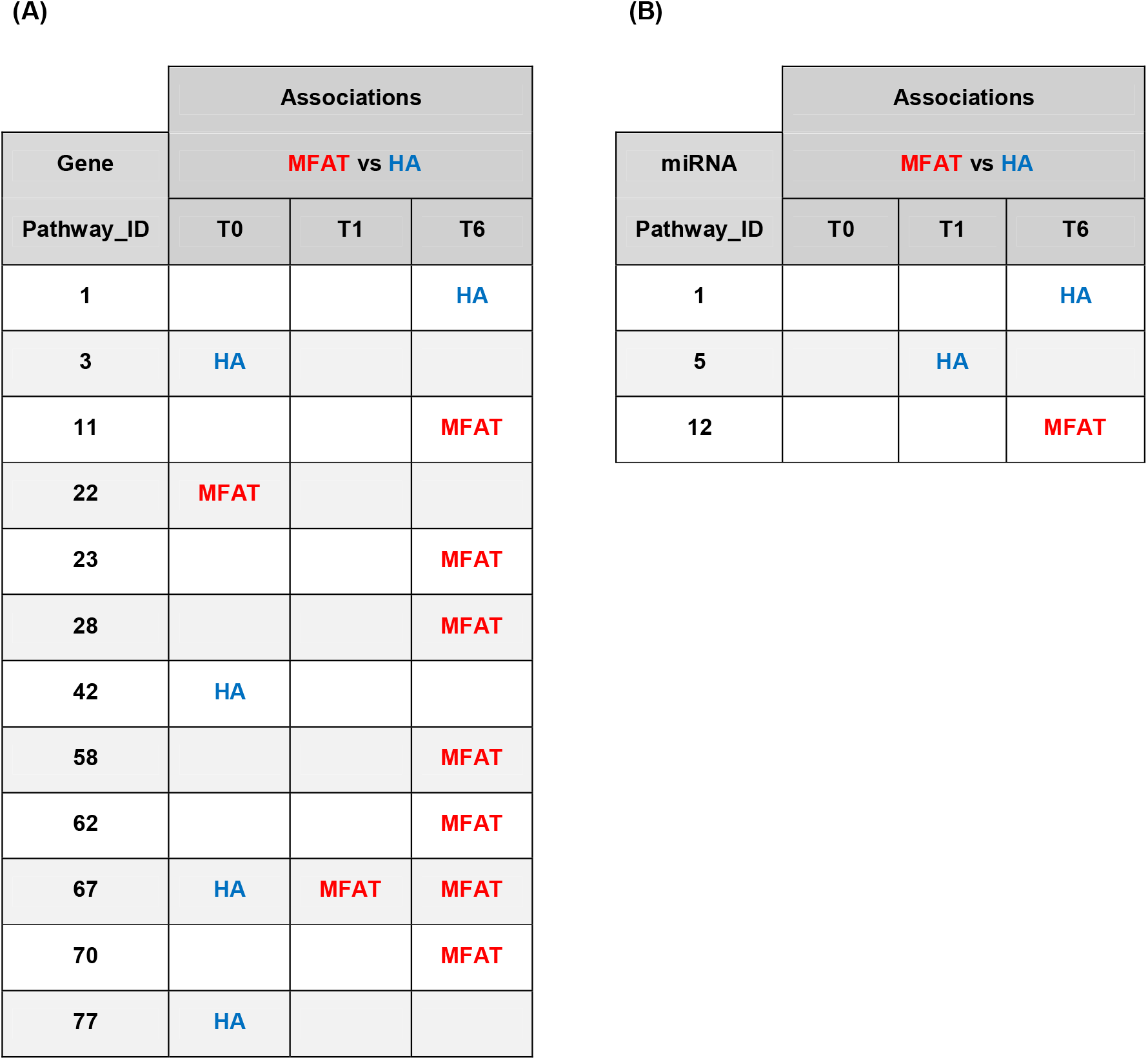
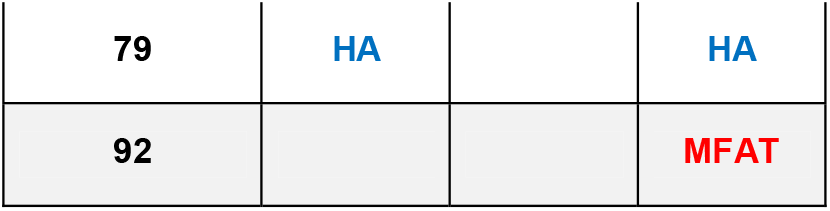
Preferential associations of the data-driven pathways. For the pathways of genes (A) or of miRNAs (B), the tables indicate if a pathway (rows) is significantly preferentially associated with MFAT-treated patients or with HA-treated patients at a given timepoint (columns). Due to space limitations, the pathways with no significant preferential association are not reported.

| (A) |  |  |  | (B) |  |  |  |
|---|---|---|---|---|---|---|---|
|  | Associations |  |  |  | Associations |  |  |
| Gene | MFAT vs HA |  |  | miRNA | MFAT vs HA |  |  |
| Pathway_ID | T0 | T1 | T6 | Pathway_ID | T0 | T1 | T6 |
| 1 |  |  | HA | 1 |  |  | HA |
| 3 | HA |  |  | 5 |  | HA |  |
| 11 |  |  | MFAT | 12 |  |  | MFAT |
| 22 | MFAT |  |  |  |  |  |  |
| 23 |  |  | MFAT |  |  |  |  |
| 28 |  |  | MFAT |  |  |  |  |
| 42 | HA |  |  |  |  |  |  |
| 58 |  |  | MFAT |  |  |  |  |
| 62 |  |  | MFAT |  |  |  |  |
| 67 | HA | MFAT | MFAT |  |  |  |  |
| 70 |  |  | MFAT |  |  |  |  |
| 77 | HA |  |  |  |  |  |  |
| 79 | HA |  | HA |  |  |  |  |
| 92 |  |  | MFAT |  |  |  |  |

For these data-driven pathways, we summarized their enriched GO-BP annotations using the REVIGO web server. By focusing on the five gene data-driven pathways with enriched GO-BP annotations that are relevant to KOA (namely gene pathways 1, 62, 70, 79 and 92), we observe that six months after treatments, the HA-treated patients remain preferentially associated with biological processes involved in KOA pathogenesis, including “protein O-linked glycosylation” and “serotonin receptor signalling” (gene pathway 1, Supplementary table 1), as well as “TOR signalling” and “histamine secretion” (gene pathway 79, Supplementary Table 9). Notably, these pathways are involved in cartilage degradation, inflammation and pain signalling.

On the other hand, the MFAT patients show preferential associations with biological processes that are involved in tissue regeneration, such as “canonical Wnt signaling pathway” and “interleukin-10-mediated signaling pathway” (gene pathway 62, Supplementary Table 6), “positive regulation of bone mineralization” and the “positive regulation of osteoblast differentiation” (gene pathway 70, Supplementary Table 8). In addition, MFAT-treated patients show preferential association with processes involved in the resolution of inflammation, with biological process such as “negative regulation of type 1 interferon production” (gene pathway 92, Supplementary Table 10), and also with the negative regulation of processes that are known to drive osteoarthritis, such as “negative regulation of activin” (gene pathway 62, Supplementary Table 6) and “negative regulation of activin receptor signaling pathway” (gene pathway 70, Supplementary Table 8).

The miRNA pathways mirror these findings. The HA-treated patients remain preferentially associated with “pseudopodium organization” (miRNA pathway 1, see Supplementary Table 11), which is linked to inflammatory OA invasiveness. On the other hand, the MFAT-treated patients are preferentially associated with regenerative and cell-adhesion processes, such as “canonical Wnt signaling pathway”, “regulation of bone mineralization” and “cell adhesion mediated by integrin” (miRNA pathway 12, Supplementary Table 12). In the next section, we further characterize the molecular signature of the KOA treatments by investigating the genes and miRNAs inside our data-driven pathways that drive their preferential associations to either MFAT-treated or HA-treated patients.

## 3. DISCUSSION

The central finding of this study is that, six months after a single intra-articular injection, MFAT and HA leave qualitatively different molecular footprints in the circulation: HA-treated patients remain associated with canonical osteoarthritis pathogenesis, whereas MFAT-treated patients shift toward chondroprotection, resolution of inflammation and bone–cartilage homeostasis. This is, to our knowledge, the first in vivo molecular correlate in humans of a regenerative therapy acting on disease biology rather than on symptoms. Below, we trace this divergence to its gene- and miRNA-level drivers and consider its translational implications.

### 3.1. Gene- and miRNA-level signatures of MFAT and HA treatments

To prioritize the genes and the miRNAs that drive the preferential associations of our pathways to either the MFAT-treated or the HA-treated patients at timepoint T6, we exploited the data-driven patient–gene association scores from the reconstructed matrix *R_4_* and the patient–miRNA association scores from the reconstructed matrix R_5_. We directly compared the average association score between a macro-molecule (gene or miRNA) and all the MFAT-treated patients against the average association score between the macro-molecule and all the HA-treated patients. Intuitively, when the ratio between these two values is greater than one, the larger the value, the more the macro-molecule is preferentially associated with the MFAT-treated patients. In the same vein, when the ratio is smaller than one, the lower the value, the more the macro-molecule is preferentially associated with the HA-treated patients. Using this approach, significant preferential associations were discovered for both HA-treated and MFAT-treated patients, revealing genes and miRNA directly or indirectly related to the molecular processes involved in OA pathogenesis (Figure 3).

**Figure 3.**
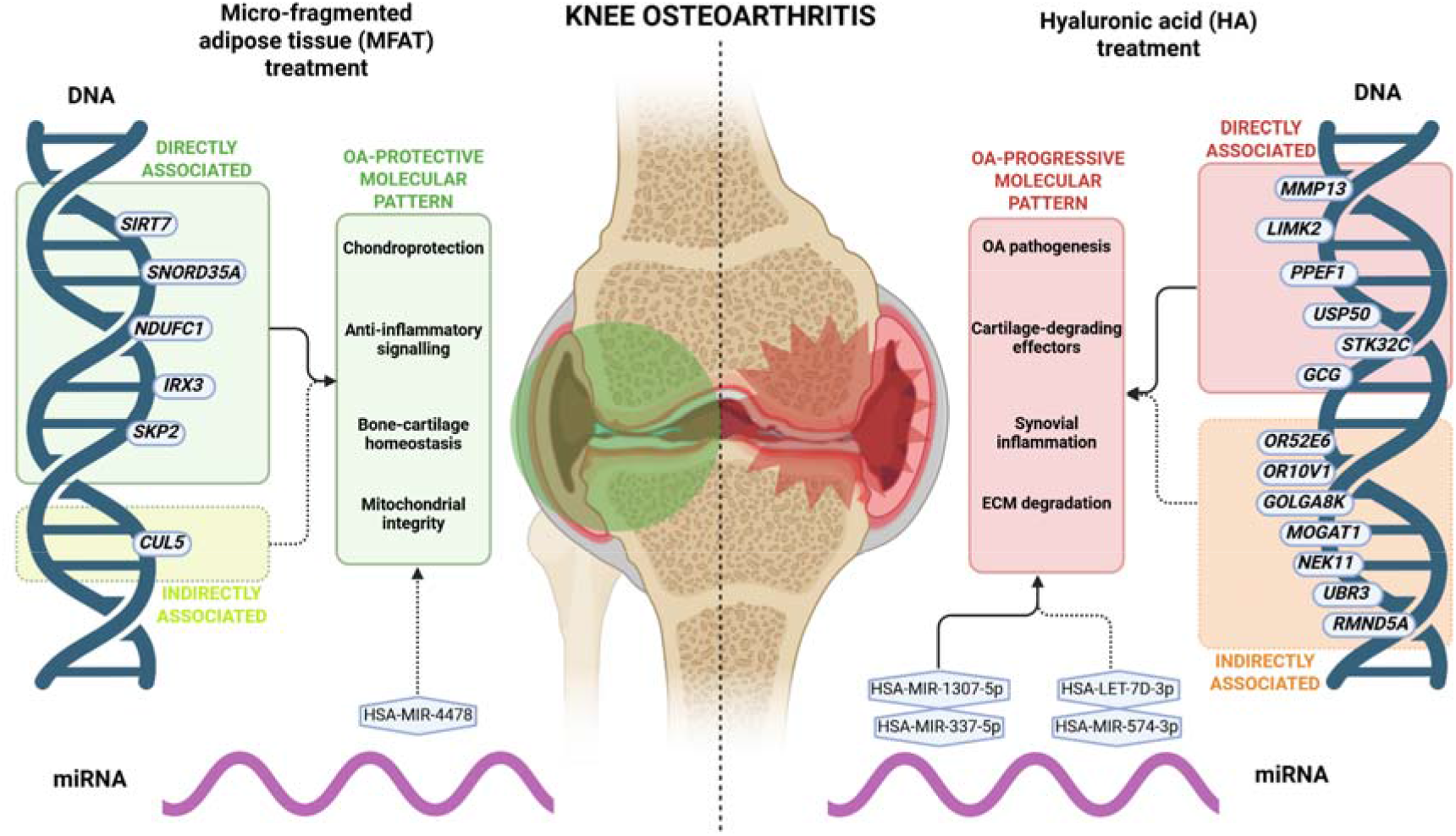
Visual summary of identified genes and miRNA preferentially associated with MFAT-treated (left) or HA-treated (right) patients, involved in OA-pathogenesis. Overall, an OA-protective molecular pattern was observed in clusters preferentially associated with MFAT-treated patients, while an OA-progressive molecular pattern was observed in clusters preferentially associated with HA-treated patients (created with Biorender.com).

For the two data-driven pathways of genes that are preferentially associated with the HA-treated patients (gene pathways 1 and 79), the top-10 prioritized genes in them are largely supported by the literature, with 13 out of 20 prioritized genes having literature evidence of their role in OA (see Supplementary Tables 13 and 16). For instance, in gene pathway 1, four of the top-10 prioritized genes are known in the literature to be involved in OA (see details in Supplementary Table 13). Gene *LIMK2* is a known prognostic indicator of OA ^18^, and studies suggest that silencing *LIMK2* can reduce osteoblasts’ mechanosensitivity ^19^. Also, gene *PPEF1* is a known biomarker of OA that is significantly upregulated in severe OA cases ^20^. In addition, variants of gene *USP50* are known pathogenetic markers of OA ^21^, and finally, gene *STK32C* is part of the epigenetic signature of OA ^22^. Furthermore, five of the remaining genes can be indirectly linked with OA through their gene families or through their biological functions. Genes *OR52E6* and *OR10V1* both code for G Protein-Coupled Receptor (GPCR) proteins, and GPCRs are known to play critical roles in OA pathogenesis ^23^. Gene *GOLGA8K* codes for a golgin protein, and the dysregulation of golgin proteins is involved in various bone diseases, including OA ^24^. Gene *MOGAT1* is involved in triglyceride synthesis, and an elevated level of triglycerides is a marker and may be involved in the pathogenesis of arthritis ^25^. Finally, gene *NEK11* is involved in inflammation, and another *NEK* gene has been shown to suppress autophagy and to intensify joint inflammation in OA ^26^.

Gene pathway 79 shows similar patterns. Two of its top-10 prioritized genes have been directly related to OA in the literature: gene *GCG* is a Pro-Glucacon gene (precursor of GLP-1) involved in the pathogenesis of OA ^27^, and gene *MMP13* is a known driver of cartilage degradation in OA ^28^. Two other genes can be indirectly linked with OA: genes *UBR3* and *RMND5A* are E3 Ubiquitin ligases, a family of genes that is known to affect OA by regulating the degradation of extracellular matrix and the inflammatory response ^29^.

On the other hand, the prioritized genes for the pathways that are preferentially associated with MFAT-treated patients (namely gene pathways 62, 70 and 92) are less supported in the literature, with only 6 out of the 30 prioritized genes having evidence of their role in OA (see Supplementary Tables 14, 15 and 17 for details). For gene pathway 62, two of the top-10 prioritized genes are known to be relevant to OA: gene *SIRT7* is known to protect against chondrocyte degeneration in OA ^30^, and gene *SNORD35A* is a known marker of OA after injury ^31^. In gene pathway 70, two of the top-10 prioritized genes are known to be relevant to OA: gene *IRX3* is known to play a role in the comorbidity between obesity and OA ^32^, and the downregulation of gene *NDUFC1* is known to lead to mitochondrial dysfunction, collapse in chondrocytes and the suppression of osteoblast differentiation ^33,34^. Finally, in gene pathway 92, one of the top-10 prioritized genes has a role in OA: gene *SKP2* has pro-inflammatory and catabolic roles in OA ^35^. One additional gene may have a role in OA: *CUL5* forms part of an E3 ubiquitin ligase complex, and E3 Ubiquitin ligases are known to regulate the degradation of the extracellular matrix and the inflammatory response in OA ^29^.

Interestingly, the prioritized miRNAs from miRNA pathway 1, which is preferentially associated with the HA-treated patients, show larger support for their role in OA than the prioritized miRNAs from miRNA pathway 12, which is preferentially associated with the MFAT-treated patients. For miRNA pathway 1 (Supplementary Table 18), two out of the top-10 prioritized miRNAs are directly related to OA in MirBase ^36^: HSA_MIR_337_5P and HSA_MIR_1307_5P. Two additional miRNAs may be related to OA due to their reported involvement in rheumatoid arthritis in MirBase: HSA_LET_7D_3P and HSA_MIR_574_3P. On the other hand, for miRNA pathway 12 (Supplementary Table 19), only one miRNA may be related to OA due to its reported involvement in rheumatoid arthritis in MirBase, namely HSA_MIR_4478. While our results on miRNAs call for further biological validation of the prioritized miRNAs, our results on genes suggest that after treatment, HA-treated patients remain preferentially associated with OA pathogenesis and cartilage-degrading effectors, such as *MMP13*, *LIMK2*, *PPEF1* and *UBR3*, while MFAT-treated patients are preferentially associated with chondroprotective effectors, including *SIRT7* and *NDUFC1*.

### 3.2. Translational and clinical implications

The findings derived from our data-fusion network carry several implications for the management of inflammatory-phenotype KOA. Most directly, they offer the first in vivo molecular rationale for the differential clinical efficacy observed in our trial. The persistence of canonical OA pathogenic processes in HA-treated patients, including cartilage-degrading effectors such as *MMP13* and *LIMK2* and markers of synovial inflammation, is notable because cartilage-specific deletion or pharmacological inhibition of *MMP13* decelerates the OA-like phenotype in surgically induced models, indicating that *MMP13* inhibition is a candidate therapeutic strategy for OA. MMP-13 is the primary matrix metalloproteinase involved in cartilage degradation through its ability to cleave type II collagen, which makes it an attractive therapeutic target, so a treatment that leaves this axis active is, in molecular terms, leaving the core driver of structural progression untouched ^28^. By contrast, the shift toward chondroprotection, anti-inflammatory signalling and bone–cartilage homeostasis in MFAT-treated patients is consistent with the proposed mode of action of adipose-derived therapy, in which the regenerative and anti-inflammatory potential of mesenchymal stromal cells, acting through immunomodulatory, anti-inflammatory and angiogenic bioactive molecules and chondrogenic differentiation, drives interest in this approach ^37^. Together, this pattern is consistent with MFAT acting on disease biology rather than on symptoms alone.

This distinction matters because the central unmet need in KOA is structural disease modification rather than further symptomatic relief, and current treatments primarily alleviate symptoms but fail to halt disease progression, underscoring the need for disease-modifying therapies. It is also clinically relevant to the standing of HA itself. Although HA produced meaningful symptomatic benefit in our trial, its place in guidelines remains contested: most guidelines conditionally recommend against intra-articular hyaluronic acid for knee osteoarthritis, reflecting a large evidence-to-practice gap, and consensus efforts have responded by trying to define which patient characteristics and phenotypes are most likely to benefit from viscosupplementation ^38^. A molecular readout that captures the residual pathogenic activity left by HA could help sharpen exactly this kind of patient-level decision.

The work also illustrates a translational strategy that could generalise beyond this trial. Because the signatures were recovered from plasma rather than from joint tissue, they are accessible through routine blood sampling, which is an important practical advantage, since synovial fluid collection is restricted in clinical practice owing to the risks associated with needle insertion into the joint space, making minimally invasive systemic biospecimens such as plasma highly advantageous for indicating OA severity and progression ^39^. This aligns with a broader move in the field toward molecular endotyping: blood-based tissue-turnover biomarker panels can identify OA endotypes that remain stable within a clinical trial and enable matching of patients to a drug’s mode of action, and more generally, biomarkers can serve as tools for patient stratification in trials, for disease management, and for directing more rational and targeted OA drug development. The interpretable, pathway-level output of our data-fusion framework is well suited to this purpose, because it yields specific genes and miRNAs that could be developed into a focused panel rather than an opaque score, and could in principle stratify patients by predicted response to support more rational selection between MFAT, HA and other interventions ^40^. This is a particularly relevant goal for a resource-intensive cell-based therapy such as MFAT, where directing treatment to the patients most likely to benefit has clear clinical and economic value.

These implications should be read as prospects rather than established clinical tools. The candidate markers were prioritised from a small cohort and from a systemic readout. Realising the clinical potential outlined here will require validation of the prioritised markers in larger cohorts, confirmation that the plasma signatures track joint-level disease activity, and ultimately linkage of these early molecular changes to structural and patient-reported endpoints over extended follow-up.

### 3.3. Strengths and limitations

To our knowledge, this is the first study to directly compare the in vivo molecular response to MFAT and HA in human inflammatory-phenotype KOA, a gap previously bridged only by in vitro assays and narrow biomarker panels. The data derive from a double-blind randomized cohort (ISRCTN88966184) sampled longitudinally at baseline, one and six months, linking the observed molecular differences to a controlled treatment contrast rather than to cross-sectional confounding. Rather than examining single cytokines or isolated miRNAs, we integrated five patient-derived omics layers, including proteomics, N-glycomics, miRNA transcriptomics, copy number variation and coding mutations, with prior protein–protein and miRNA–gene regulatory networks in a single data-fusion model, preserving relational structure that naïve concatenation would discard. The NMTF framework is also interpretable by construction: it yields data-driven gene and miRNA pathways and explicit patient–gene and patient–miRNA associations, allowing each treatment’s signature to be traced to specific functional modules and their driver molecules. Finally, the pathway-level analysis aggregates weak individual effects into coherent modules, enhancing power where single-molecule signals are too small to detect, and the biological coherence of these modules is supported by their significant enrichment in independent GO-BP and Reactome Pathway annotations.

Where limitations are concerned, the cohort is small (p = 41), typical of translational trials but limiting statistical power and subgroup resolution; the results are therefore hypothesis-generating and require validation in a larger, independent cohort. The analysis is based on plasma, a systemic but indirect readout of a disease whose proximal drivers reside in the joint and synovium, so the degree to which the circulating signatures reflect local joint biology remains unclear. The miRNA pathways were annotated by proxy via their MIRNET target genes, inheriting any bias in the underlying database, and the prior networks we used (BioGRID, MIRNET) are themselves incomplete and skewed toward well-studied genes. Methodologically, the number of clusters was fixed by a rule of thumb rather than optimized, and k-means is stochastic. We mitigated this by selecting the most representative of 25 clusterings, but the pathway boundaries are not unique. Driver genes and miRNAs were prioritized by a ratio of mean association scores rather than formal testing, since features far outnumber patients, so these rankings are best read as candidates for follow-up. Finally, the signatures we report are statistical associations. Confirming that MFAT’s chondroprotective, anti-inflammatory shift is causal and translates into long-term structural disease modification will require functional validation and extended follow-up.

### 3.4. Future perspectives and conclusions

The findings reported in this study open several avenues for follow-up. The most immediate is validation: the divergent molecular signatures of MFAT and HA should be tested in a larger, independent cohort, ideally one powered for the patient subgroups that current sample sizes cannot resolve. Because our readout is systemic, a natural next step is to determine how faithfully the circulating signatures reflect events in the joint itself, for example, by relating them to synovial or synovial-fluid measurements where paired samples are available. The pathways and driver molecules prioritized by our framework also provide concrete, testable hypotheses: the chondroprotective, anti-inflammatory and bone–cartilage homeostatic processes attributed to MFAT, and the specific genes and miRNAs ranked within those modules, are candidates for targeted functional assays that could establish causality rather than association. Methodologically, the data-fusion approach extends readily to additional molecular layers, to longer-term timepoints that would link these early signatures to structural outcomes, and in principle to other inflammatory-phenotype joint diseases. In the longer term, a molecular signature that distinguishes responders from non-responders could support rational patient selection and strengthen the regulatory case for MFAT as a disease-modifying intervention.

In conclusion, we applied an interpretable multi-omics data-fusion framework to longitudinally collected plasma from a double-blind randomized trial in inflammatory-phenotype KOA, integrating proteomics, N-glycomics, miRNA transcriptomics and whole-genome sequencing data with prior molecular networks across three timepoints. By six months, the two treatments left clearly distinct molecular signatures: HA was dominated by canonical OA pathogenic processes, including cartilage-degrading effectors and synovial inflammation, whereas MFAT shifted the systemic landscape toward chondroprotection, anti-inflammatory signalling and bone–cartilage homeostasis. To our knowledge, these are the first systems-level molecular data to directly compare the in vivo response to the two treatments in human KOA, and they provide initial evidence that MFAT acts as a disease-modifying intervention. While these results are hypothesis-generating and require validation, they demonstrate the value of interpretable data fusion for uncovering treatment mechanisms in small translational cohorts and offer a molecular foundation for the rational use of MFAT in inflammatory KOA.

## 5. METHODS

### 5.1. Clinical recruitment, phenotyping, randomization and intervention

The present work is a retrospective multi-omics analysis of biospecimens collected from participants of a completed prospective, randomized, double-blind, active-controlled clinical trial (ISRCTN88966184), the design, conduct, and clinical and imaging outcomes of which have been reported in full elsewhere ^8^. The parent trial was carried out between February 2020 and May 2023 within project KK.01.2.1.02.0173, coordinated by St. Catherine Specialty Hospital (Zagreb, Croatia) in collaboration with Clinical Hospital Centre Sestre Milosrdnice and University Hospital Merkur, in accordance with the Declaration of Helsinki, Good Clinical Practice, and the EU General Data Protection Regulation. The trial protocol was approved by the institutional review boards of all three participating centres (St. Catherine Specialty Hospital 22/5-I; University Hospital Merkur BR-0311-2758; University Hospital Centre Sestre Milosrdnice 251-29-11/3-22-02), and all participants provided written informed consent at enrolment, including consent for the use of their biospecimens in subsequent molecular analyses. The subsequent molecular analyses were also approved by the institutional review board at which they were carried out (St. Catherine Specialty Hospital 25/2-I).

For context, the parent trial enrolled adults aged 30–75 years with primary knee osteoarthritis fulfilling Dell’Isola inflammatory-phenotype criteria ^3^, including a MOAKS synovitis/effusion score of 2 or 3, who were randomized 2:1 to a single intra-articular injection of autologous micro-fragmented adipose tissue (MFAT) prepared with the Lipogems® Ortho Kit (Lipogems International SpA, Milan, Italy) or 60 mg of high-molecular-weight hyaluronic acid (Hyalubrix 60®, Fidia S.p.A., Padova, Italy), delivered under ultrasound guidance. All participants underwent abdominal lipoaspiration regardless of allocation to preserve blinding. Clinical response was assessed at T0, T1, and T6 using the KOOS, WOMAC, and VAS instruments; structural response was assessed at T0 and T6 by dGEMRIC with T1 mapping in seven cartilage regions of interest on a Siemens MAGNETOM® Lumina 3T scanner. Responder definitions used in the present analysis follow the published protocol: KOOS Pain ≥10-point improvement, WOMAC Total ≥15-point improvement, VAS Movement reduction >2 points, and dGEMRIC structural response defined as a ≥10% increase in ≥3 of 7 cartilage regions ^8^.

### 5.2. Multi-omics generation - proteomics, glycomics, miRNA, and genomics

Peripheral venous blood was collected from all enrolled patients at baseline (T0), one month (T1), and six months (T6) post-intervention for plasma proteomic, N-glycomic, and miRNA transcriptomic profiling. Plasma was separated and stored at −80 °C at the Tissue and Cell Bank of the Department of Traumatology, Clinical Hospital Centre Sestre Milosrdnice, and shipped on dry ice under temperature-monitored conditions to the analytical laboratories. Germline whole-genome sequencing was performed using DNA obtained from a dedicated post-trial blood draw. Of the 53 enrolled patients, 41 responded to the recall and provided samples for whole-genome sequencing, constituting the multi-omics analysis cohort.

Genomic DNA was extracted from peripheral leukocytes using the Mag-Bind Blood & Tissue DNA HDQ Kit (Omega Bio-Tek, Norcross, GA, USA). DNA quality and quantity were assessed by Qubit 1X dsDNA HS Assay (Life Technologies, Eugene, OR, USA), NanoDrop spectrophotometry, and Agilent Genomic DNA ScreenTape and High-Sensitivity D1000 ScreenTape assays (Agilent Technologies, Waldbronn, Germany). Sequencing libraries were prepared with the Illumina DNA Prep (M) Tagmentation Kit and sequenced on the Illumina NovaSeq 6000 platform (Illumina, San Diego, CA, USA) using 2 × 150 bp paired-end chemistry to a mean coverage of approximately 100×. Primary and secondary analyses were performed on the Illumina DRAGEN platform following the GATK best-practices pipeline, including Variant Quality Score Recalibration. Sequence reads were aligned to the GRCh37/hg19 reference genome using BWA. Single-nucleotide variants and small insertions/deletions were annotated against ClinVar, OMIM, the GWAS Catalog, HGMD, and SwissVar, and filtered against allele-frequency data from 1000 Genomes Phase 3, ExAC, EVS, dbSNP147, and an internal population database. Functional impact of non-synonymous variants was predicted using PolyPhen-2, SIFT, MutationTaster2, MutationAssessor, and LRT, and variants were classified according to ACMG/AMP guidelines. The resulting per-patient variant matrix used as input to data fusion comprised 18,886 genes carrying SNVs/InDels across the 41 patients.

Plasma concentrations of inflammatory, metabolic, and matrix-remodelling mediators were quantified using a custom multiplex sandwich immunoassay designed for this study and run on the Bio-Plex® 200 system (Bio-Rad Laboratories, Hercules, CA, USA) at Labena d.o.o. (Ljubljana, Slovenia). The panel targeted 40 analytes encoded by 39 genes: IL-1β, IL-1Ra, IL-2, IL-4, IL-5, IL-6, IL-8, IL-10, IL-12, IL-13, IL-15, IL-17A, IL-18, IL-21, IL-22, TNF-α, IFN-γ, TGF-β1, TGF-β2, MMP-1, MMP-2, MMP-3, MMP-8, MMP-9, MMP-13, TIMP-1, TIMP-2, TIMP-4, sICAM-1, V-CAM-1, VEGF, leptin, CCL2 (MCP-1), CCL3 (MIP-1α), CCL4 (MIP-1β), CCL5 (RANTES), CCL7 (MCP-3), and CXCL-12 (SDF-1). To minimise batch effects, samples were processed in groups containing all timepoints for both treatment arms in each plate. IL-4 and IL-5 yielded insufficient signal across the cohort and were excluded from downstream data fusion, yielding a final 38-protein input matrix.

Plasma N-glycome profiling was performed at Genos Glycoscience Research Laboratory (Zagreb, Croatia). Plasma aliquots were randomized across 96-well plates with standard samples included in tetraplicate for quality control. For each sample, 10 μL of plasma was denatured with 20 μL of 2% (w/v) sodium dodecyl sulfate (Invitrogen, Carlsbad, CA, USA) at 65 °C for 10 min, after which 10 μL of 4% (v/v) Igepal CA-630 (Sigma-Aldrich, St. Louis, MO, USA) was added and the mixture agitated for 15 min. N-glycans were released by overnight incubation (18 h) with 1.2 U of PNGase F (Promega, Madison, WI, USA) at 37 °C and fluorescently labelled by reductive amination with 2-aminobenzamide using 2-picoline borane (both Sigma-Aldrich) as the reducing agent at 65 °C for 2 h. Labelled glycans were purified by hydrophilic interaction solid-phase extraction on a 0.2 μm wwPTFE 96-well filter plate (Pall Corporation, Port Washington, NY, USA) and eluted in 180 μL of ultrapure water. Separation was performed on a Waters ACQUITY H-Class UHPLC system using a 150 mm ACQUITY Premier Column maintained at 10 °C, with a linear gradient from 30% to 70% acetonitrile in 100 mM ammonium formate (pH 4.4) over 32.5 min at 0.561 mL/min and fluorescence detection at excitation 250 nm/emission 428 nm using ACQUITY Premier FLR detectors (Waters, Milford, MA, USA). Chromatograms were manually integrated using Empower 3 software (Waters), and each peak was expressed as a percentage of the total integrated area. After quality control filtering across all samples and timepoints, 33 N-glycan features were retained as the input matrix for data fusion.

Small RNA sequencing was performed at TAmiRNA GmbH (Vienna, Austria), a CRO specialising in cell-free RNA biomarker discovery. Small RNA-seq libraries were prepared from plasma-derived total RNA and sequenced on an Illumina platform following TAmiRNA’s standard small RNA-seq workflow, with miND® spike-in controls included in each library for inter-sample normalisation and quality control ^41^. Raw sequencing reads were processed using the miND® (miRNA NGS Discovery) pipeline ^42^, a Snakemake-based workflow that performs adapter and quality trimming, FastQC- and multiQC-based quality control, and hierarchical alignment with bowtie1 against the human reference genome (Ensembl release 105) and miRDeep2-based quantification of mature miRNAs against miRBase v22.1, with additional small RNA classification against RNAcentral. miRNAs were quantified as reads per million mapped miRNAs, and low-abundance features were removed using the independent-filtering procedure implemented in miND. Of the 1,947 mature miRNAs detected across the cohort, 1,890 were retained after intersection with the MIRNET miRNA–gene regulatory database (v2.0) and used as the input matrix for data fusion.

### 5.3. Data fusion network (NMTF)

To holistically mine the multi-omics KOA data and enable a data-driven pathway-level analysis of the molecular differences between MFAT and HA treatments, we fused the multi-omics data from the patients with the experimentally validated physical interactions between the genes’ products from the BioGRID database ^43^, and the miRNA–gene regulatory interactions from the MIRNET database ^44^, by using non-negative matrix tri-factorization (NMTF), a co-clustering, dimensionality-reduction and inference method ^16^. To enable their fusion with NMTF, we modeled the multi-omics data of a given timepoint as seven data matrices: matrix *A_1_* captured the physical interactions between the genes’ products (from BioGRID database), *A_2_*captured the copy number variation profiles (CNVs) of the patients, *A_3_*captured the single nucleotide polymorphisms and insertion/deletions profiles (SNPIDs) of the patients, *A_4_* captured the proteomics profiles of the patients, *A_5_* captured the miRNA transcriptomics profiles of the patients, *A_6_* captured the glycomics profiles of the patients, and *A_7_* captured the regulatory interactions between the miRNAs and the genes (from MIRNET database) (Figure 4).

**Figure 4.**
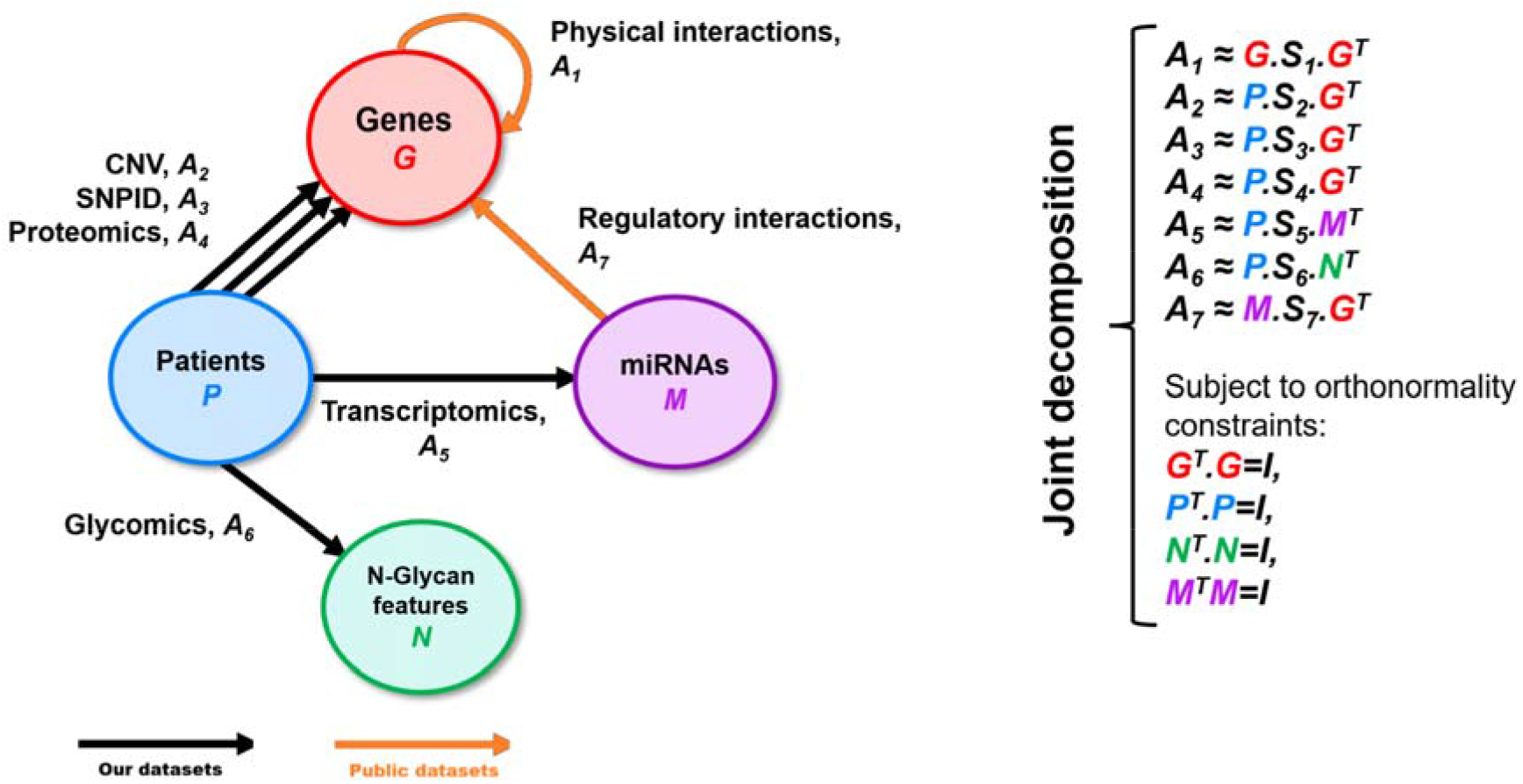
Overview of our timepoint-specific data-fusion methodology. For each timepoint, the multi-omics data between the patients, genes, miRNAs and N-glycans features are modeled as seven input data matrices, *A_1_* to *A_7_*. In the illustration of our data model (on the left), each input data matrix is represented by an arrow whose tail indicates the types of entities represented by its row and whose head indicates the type of entities represented by its column. As illustrated on the right, we integrate these seven data matrices by jointly decomposing them as three lower-dimensional matrix factors while sharing common matrix factors along the decompositions, which allows for learning from all the data (e.g., matrix factor *P*, which features the low-dimensional representation of the patients, is shared across the decompositions of *A_2_*, *A_3_*, *A_4_*, *A_5_* and *A_6_*). Furthermore, to reduce the dependencies in the learnt low-dimensional representations of the patients, genes, miRNAs and N-glycan features, we constrain matrix factors *P*, *G*, *M* and *N* to be orthonormal.

#### 5.3.1. Data modeling

We restricted the analysis to the *p* = 41 patients having all data modalities, *g* = 17,569 genes having both BioGRID interactions and copy number variations measured from our plasma tissue samples, *m* = 1,890 miRNAs having both MIRNET interactions with our selected genes and transcription levels measured from our plasma tissue samples, and *n* = 33 N-glycan features measured on our plasma tissues. The BioGRID physical interaction network (v5.0.252) was represented by its adjacency matrix and then transformed into its Positive Pointwise Mutual Information (PPMI) representation, *A_1_*, which captured how frequently two genes co-occur in a random walk relative to chance; the additional topological information encoded in PPMI matrices has been shown to yield embedding vectors that better capture functional relationships than the adjacency matrix alone. The CNV profiles (*A_2_*) recorded the number of copies of each gene per patient, and the SNPID profiles (*A_3_*) indicated whether a patient carries one or more coding mutations in each gene. For the proteomics data, the measured protein abundances were sparse, so we applied a network propagation pre-processing step that smooths the abundance values by spreading them over neighbouring genes in the physical interaction network, yielding matrix *A_4_*. The miRNA transcriptomics (*A_5_*) and glycomics (*A_6_*) profiles recorded the measured transcription value of each miRNA and the value of each N-glycan feature per patient, respectively, and the miRNA–gene regulatory interactions from MIRNET (v2.0) were captured as the binary matrix *A_7_*. Each timepoint is thus represented by seven data matrices, *A_1_* to *A_7_*, capturing complementary aspects of the disease biology.

#### 5.3.2. Timepoint-specific data fusion

To fuse the multi-omics data of a given timepoint, we used NMTF to jointly decompose the corresponding seven data matrices as the product of three lower-dimensional matrix factors: *A_1_* ≈ GS*_1_G^T^, A_2_*≈ PS*_2_G^T^, A_3_* ≈ PS*_3_G^T^, A_4_* ≈ PS*_4_G^T^, A_5_* ≈ PS*_5_M^T^, A_6_* ≈ PS*_6_N^T^ and A_7_* ≈ MS*_7_G^T^*(Figure 4). The key was in sharing common matrix factors across the decompositions, which allowed for learning from all the data. After the decomposition, matrix factor *G*, the cluster indicator of the genes, featured data-driven low-dimensional representations (also called embedding vectors) of the genes (the row-vectors of *G*), which indicated how the genes functionally cluster together according to all the data (i.e., functionally similar genes are assigned similar embedding vectors in *G*). Matrix factors *P*, *M* and *N* were the same, but for the patients, the miRNAs and the N-glycan features, respectively. The matrix factors *S_1_* to *S_7_* are the compressed representations of matrices *A_1_* to *A_7_*, respectively, indicating how the clusters of patients, genes, miRNAs and N-glycan features relate to each other. To reduce the dependencies in the learnt low-dimensional representations, we constrained matrix factors *P, G, M* and *N* to be orthonormal. Importantly, the reconstructed matrix *R_4_ = PS_4_G^T^* contains new, data-driven association scores between the patients and the genes that emerged from the fusion of all the data (so-called matrix completion property of NMTF). Similarly, the reconstructed matrix *R_5_* = *PS_5_M^T^* contains new, data-driven association scores between the patients and the miRNAs.

#### 5.3.3. Solving the data-fusion model

For a given timepoint, solving the joint decomposition amounted to minimising the reconstruction error across all seven matrices subject to non-negativity of the factors. As this is an NP-hard continuous optimization problem, we solved it heuristically using a fixed-point solver based on multiplicative update rules, initialised from a singular value decomposition, which converges towards a locally optimal solution. The numbers of clusters of patients, genes, miRNAs and N-glycan features were set using the standard rule of thumb, giving approximately 4, 93, 30 and 3 clusters, respectively. Applying this framework separately to each timepoint yielded timepoint-specific matrix factors.

#### 5.3.4. Uncovering data-driven pathways across timepoints

We exploited the embedding vectors of the genes and of the miRNAs to uncover data-driven pathways (clusters) of genes and of miRNAs, enabling higher-level molecular analysis of MFAT and HA treatments. Because each gene was represented by a separate embedding vector at each of the three timepoints, we obtained a single global representation per gene by concatenating its three timepoint-specific embedding vectors, and then clustered genes by applying k-means using the Euclidean distance between these global vectors. To account for the stochasticity of k-means, we repeated the clustering 25 times and retained the most representative partition, defined as the one most similar to all other runs as measured by the Rand index. The same procedure applied to the miRNA cluster indicator matrices yielded the data-driven pathways of miRNAs that account for all data and all timepoints.

#### 5.3.5. Assessing biological relevance

To assess the biological relevance of the resulting pathways, we tested whether they are statistically significantly enriched in Gene Ontology Biological Process (GO-BP) ^45^, or Reactome Pathway (RP) annotations ^46^ (both collected on 17 December 2025), using a hypergeometric test. A pathway was considered significantly enriched when its enrichment p-value, after Benjamini–Hochberg correction for multiple hypothesis testing, was lower than or equal to 5%. To enable the same analysis for the miRNA pathways, we annotated each miRNA using its target genes as a proxy, i.e., with the GO-BP and RP annotations of the genes with which it has regulatory interactions according to MIRNET. We reported both the percentage of pathways containing at least one enriched annotation and the percentage of annotations enriched in at least one pathway, and we used the REVIGO web server to summarise the enriched GO-BP terms for selected pathways ^47^.

#### 5.3.6. Discovery of preferential treatment associations

By using the data-driven association scores between the patients and the genes and between the patients and the miRNAs, we uncovered the data-driven pathways that are preferentially associated with either the MFAT-treated or the HA-treated patients after treatments (at timepoint T6) and, within them, the genes and miRNAs that drive these preferential associations, revealing the distinct molecular signatures of the two treatments. To determine whether a gene pathway is preferentially associated with one patient group over the other, we compared the distribution of association scores (entries in *R_4_*) between the genes in the pathway and the first patient group with the corresponding distribution for the second patient group, using a one-sided Mann–Whitney U test and a 5% significance threshold after Benjamini–Hochberg correction. To prioritise the individual genes driving these associations, where the Mann–Whitney test was under-powered because the genes far outnumber the patients, we instead ranked genes by the ratio of their mean association score with one patient group to that with the other, where a ratio above one indicates preferential association with the first group and a ratio below one with the second. The same procedures, applied to the association scores in *R_5_*, identify the preferentially associated miRNA pathways and their driver miRNAs.

## DATA AVAILABILITY

The datasets generated and analysed during the current study are available in the figshare repository under DOI https://doi.org/10.6084/m9.figshare.33193641.

## ACKNOWLEDGEMENTS

We would like to thank the International Society for Applied Biological Sciences, the International Center for Applied Biological Research, and St. Catherine Specialty Hospital for their ongoing support.

## ETHICS STATEMENT

This study is a secondary molecular analysis of plasma samples and clinical data collected within a prospective, randomised, double-blind clinical trial registered with the ISRCTN registry (ISRCTN88966184) and reported previously (Molnar et al., Biomedicines 2025;13:2301). All methods were carried out in accordance with the relevant guidelines and regulations. The study was conducted in accordance with the Declaration of Helsinki and was approved by the Institutional Review Board of each participating institution: St. Catherine Specialty Hospital (UR 22/5-I, 25 March 2022), University Hospital Merkur (UR BR-0311-2758, 31 March 2022) and Sestre milosrdnice University Hospital Centre (251-29-11/3-22-02, 7 April 2022). Written informed consent was obtained from all participants prior to enrolment, including consent for molecular analysis of the collected biological samples. All data were anonymised and stored in a secure, password-protected database in compliance with the GDPR and Good Clinical Practice guidelines.

## AUTHOR CONTRIBUTIONS

Conceptualization - D.P., V.M., P.B., Ž.J., G.L.; Methodology - D.P., V.M., P.B., L.B.; Software - N.P., N.M.D.; Formal analysis - U.P.Z, T.K., N.P., N.M.D., G.L.; Validation - D.P., V.M., P.B., L.B., Ž.J.; Investigation - N.M.D., V.M., P.B., L.B., T.K.; Resources - U.P.Z, T.K., N.P., N.M.D., G.L., Ž.J.; Data Curation - N.M.D., V.M., P.B., L.B.; Writing: Original Draft - D.P., V.M., P.B., L.B., N.M.D.; Writing: Review & Editing - G.L., U.P.Z., T.K., Ž.J., N.P.; Visualization - L.B., P.B., N.M.D.; Supervision - D.P., G.L., N.P.; Project administration - D.P., V.M.

## CLINICAL TRIAL NUMBER

Not applicable — the present work is a retrospective secondary molecular analysis of samples and data collected in the registered trial ISRCTN88966184.

## REFERENCES

1. GBD 2021 Osteoarthritis Collaborators. Global, regional, and national burden of osteoarthritis, 1990-2020 and projections to 2050: a systematic analysis for the Global Burden of Disease Study 2021. Lancet Rheumatol 5, e508–e522 (2023).

2. Brandt, M. D., Malone, J. B. & Kean, T. J. Advances and Challenges in the Pursuit of Disease-Modifying Osteoarthritis Drugs: A Review of 2010-2024 Clinical Trials. Biomedicines 13, (2025).

3. Dell’Isola, A., Allan, R., Smith, S. L., Marreiros, S. S. P. & Steultjens, M. Identification of clinical phenotypes in knee osteoarthritis: a systematic review of the literature. BMC Musculoskelet Disord 17, 425 (2016).

4. Sanchez-Lopez, E., Coras, R., Torres, A., Lane, N. E. & Guma, M. Synovial inflammation in osteoarthritis progression. Nat Rev Rheumatol 18, 258–275 (2022).

5. Molnar, V. et al. Cytokines and Chemokines Involved in Osteoarthritis Pathogenesis. Int J Mol Sci 22, (2021).

6. Primorac, D. et al. Knee Osteoarthritis: A Review of Pathogenesis and State-Of-The-Art Non-Operative Therapeutic Considerations. Genes (Basel) 11, (2020).

7. Hudetz, D. et al. The future of cartilage repair. in Personalized Medicine in Healthcare Systems 375–411 (Springer International Publishing, Cham, 2019).

8. Molnar, V. et al. Clinical and dGEMRIC Evaluation of Microfragmented Adipose Tissue Versus Hyaluronic Acid in Inflammatory Phenotype of Knee Osteoarthritis: A Randomized Controlled Trial. Biomedicines 13, (2025).

9. Maheu, E. et al. Why we should definitely include intra-articular hyaluronic acid as a therapeutic option in the management of knee osteoarthritis: Results of an extensive critical literature review. Semin Arthritis Rheum 48, 563–572 (2019).

10. Sherman, S. L., Gudeman, A. S., Kelly, J. D., 4th, Dimeff, R. J. & Farr, J. Mechanisms of Action of Intra-articular Hyaluronic Acid Injections for Knee Osteoarthritis: A Targeted Review of the Literature. Am J Sports Med 53, 2771–2782 (2025).

11. Nam, Y. et al. Harnessing Artificial Intelligence in Multimodal Omics Data Integration: Paving the Path for the Next Frontier in Precision Medicine. Annu Rev Biomed Data Sci 7, 225–250 (2024).

12. Čopar, A., Zupan, B. & Zitnik, M. Fast optimization of non-negative matrix tri-factorization. PLoS One 14, e0217994 (2019).

13. Riccio-Rengifo, C., Cascianelli, S., Ceddia, G. & Masseroli, M. Inferring breast cancer subtype associations using an original omics integration based on non-negative matrix Tri-factorization. in Lecture Notes in Computer Science 255–272 (Springer Nature Switzerland, Cham, 2025).

14. Tang, X. et al. Indicator Regularized Non-Negative Matrix Factorization Method-Based Drug Repurposing for COVID-19. Front Immunol 11, 603615 (2020).

15. Xenos, A., Malod-Dognin, N., Zambrana, C. & Pržulj, N. Integrated Data Analysis Uncovers New COVID-19 Related Genes and Potential Drug Re-Purposing Candidates. Int J Mol Sci 24, (2023).

16. Pržulj, N. & Malod-Dognin, N. Simplicity within biological complexity. Bioinform Adv 5, vbae164 (2025).

17. Zitnik, M. et al. Current and future directions in network biology. Bioinform Adv 4, vbae099 (2024).

18. Wang, J. et al. Screening crucial lncRNAs and genes in osteoarthritis by integrated analysis. Adv Rheumatol 63, 7 (2023).

19. Yang, Z. et al. Inhibition of FSS-induced actin cytoskeleton reorganization by silencing LIMK2 gene increases the mechanosensitivity of primary osteoblasts. Bone 74, 182–190 (2015).

20. Chou, C.-H. et al. Genome-wide expression profiles of subchondral bone in osteoarthritis. Arthritis Res Ther 15, R190 (2013).

21. Zengini, E. et al. Genome-wide analyses using UK Biobank data provide insights into the genetic architecture of osteoarthritis. Nat Genet 50, 549–558 (2018).

22. Izda, V. et al. A Pilot Analysis of Genome-Wide DNA Methylation Patterns in Mouse Cartilage Reveals Overlapping Epigenetic Signatures of Aging and Osteoarthritis. ACR Open Rheumatol 4, 1004–1012 (2022).

23. Wen, Z.-Q. et al. G Protein-Coupled Receptors in Osteoarthritis: A Novel Perspective on Pathogenesis and Treatment. Front Cell Dev Biol 9, 758220 (2021).

24. Iacobescu, G. L. et al. Exploring the Implications of Golgi Apparatus Dysfunction in Bone Diseases. Cureus 16, e56982 (2024).

25. Ma, W., Chen, H., Deng, J., Yuan, Q. & Li, H. The role of triglycerides in predicting new-onset arthritis in the general population over 45 years old: evidence from the China health and retirement longitudinal study. Front Endocrinol (Lausanne) 16, 1530874 (2025).

26. Li, Z., Li, X., Xia, H., Wang, Y. & Wei, N. NEK2 promotes the progression of osteoarthritis by stabilizing ATF2 through phosphorylation at Ser-112 and inhibiting autophagy. Int Immunopharmacol 146, 113833 (2025).

27. Cheng, J., Solomon, T., Estee, M., Cicuttini, F. M. & Lim, Y. Z. Effect of glucagon-like peptide-1 receptor agonists in osteoarthritis: A systematic review of pre-clinical and human studies. Osteoarthr Cartil Open 7, 100567 (2025).

28. Hu, Q. & Ecker, M. Overview of MMP-13 as a Promising Target for the Treatment of Osteoarthritis. Int J Mol Sci 22, (2021).

29. Jin, Z. et al. The multifaceted roles of E3 ubiquitin ligases in osteoarthritis. Front Cell Dev Biol 13, 1665313 (2025).

30. Wu, S.-Y., Du, Y.-C. & Yue, C.-F. Sirt7 protects chondrocytes degeneration in osteoarthritis via autophagy activation. Eur Rev Med Pharmacol Sci 24, 9246–9255 (2020).

31. Peffers, M. J. et al. SnoRNA signatures in cartilage ageing and osteoarthritis. Sci Rep 10, 10641 (2020).

32. Arruda, A. L. et al. Genetic underpinning of the comorbidity between type 2 diabetes and osteoarthritis. Am J Hum Genet 110, 1304–1318 (2023).

33. Tao, H. et al. The Emerging Role of the Mitochondrial Respiratory Chain in Skeletal Aging. Aging Dis 15, 1784–1812 (2024).

34. Lee, S.-Y. & Long, F. Notch signaling suppresses glucose metabolism in mesenchymal progenitors to restrict osteoblast differentiation. J Clin Invest 128, 5573–5586 (2018).

35. Huang, Y., Pan, W. & Ma, J. SKP2-mediated ubiquitination and degradation of KLF11 promotes osteoarthritis via modulation of JMJD3/NOTCH1 pathway. FASEB J 38, e23640 (2024).

36. Kozomara, A., Birgaoanu, M. & Griffiths-Jones, S. miRBase: from microRNA sequences to function. Nucleic Acids Res 47, D155–D162 (2019).

37. De Groote, J. et al. Autologous Micro-Fragmented Adipose Tissue (MFAT) Injections May Be an Effective Treatment for Advanced Knee Osteoarthritis: A Longitudinal Study. J Clin Med 14, (2025).

38. Pisaniello, H. L., Goh, S. & Buchbinder, R. Intra-articular hyaluronic acid (viscosupplementation) for osteoarthritis: is it effective? Aust Prescr 49, 16–21 (2026).

39. Zhang, X. et al. Immune System-Related Plasma Pathogenic Extracellular Vesicle Subpopulations Predict Osteoarthritis Progression. Int J Mol Sci 25, (2024).

40. Mobasheri, A. et al. Molecular taxonomy of osteoarthritis for patient stratification, disease management and drug development: biochemical markers associated with emerging clinical phenotypes and molecular endotypes. Curr Opin Rheumatol 31, 80–89 (2019).

41. Khamina, K. et al. A MicroRNA Next-Generation-Sequencing Discovery Assay (miND) for Genome-Scale Analysis and Absolute Quantitation of Circulating MicroRNA Biomarkers. Int J Mol Sci 23, (2022).

42. Diendorfer, A., Khamina, K., Pultar, M. & Hackl, M. miND (miRNA NGS Discovery pipeline): a small RNA-seq analysis pipeline and report generator for microRNA biomarker discovery studies. F1000Res. 233 (2022) doi:10.12688/f1000research.94159.1.

43. Oughtred, R. et al. The BioGRID database: A comprehensive biomedical resource of curated protein, genetic, and chemical interactions. Protein Sci 30, 187–200 (2021).

44. Chang, L. & Xia, J. MicroRNA Regulatory Network Analysis Using miRNet 2.0. Methods Mol Biol 2594, 185–204 (2023).

45. Ashburner, M. et al. Gene ontology: tool for the unification of biology. The Gene Ontology Consortium. Nat Genet 25, 25–29 (2000).

46. Ragueneau, E. et al. The Reactome Knowledgebase 2026. Nucleic Acids Res 54, D673–D681 (2026).

47. Supek, F., Bošnjak, M., Škunca, N. & Šmuc, T. REVIGO summarizes and visualizes long lists of gene ontology terms. PLoS One 6, e21800 (2011).

