## Supplemental Tables for "Micro-fragmented adipose tissue reprogrammes the systemic molecular landscape of inflammatory knee osteoarthritis"

### *SUPPLEMENTARY TABLES*

| **GO Term** | **Name** |
| --- | --- |
| GO:0002477 | antigen processing and presentation of exogenous peptide antigen via MHC class Ib |
| GO:0006044 | N-acetylglucosamine metabolic process |
| GO:0006465 | signal peptide processing |
| GO:0006493 | protein O-linked glycosylation |
| GO:0006506 | GPI anchor biosynthetic process |
| GO:0050651 | dermatan sulfate proteoglycan biosynthetic process |
| GO:0007271 | synaptic transmission, cholinergic |
| GO:0007165 | signal transduction |
| GO:0095500 | acetylcholine receptor signaling pathway |
| GO:0051899 | membrane depolarization |
| GO:0032098 | regulation of appetite |
| GO:0035094 | response to nicotine |
| GO:0007210 | serotonin receptor signaling pathway |
| GO:0035095 | behavioral response to nicotine |

**Supplementary Table 1. Summary of the GO-BP terms that are enriched on gene data-driven pathway 1**, which is preferentially associated with HA-treated patients at timepoint T=6 months. We used REVIGO webserver to identify the most important GO terms (redundancy parameter =0.7). The selected GO terms are grouped together according to their semantic similarity (as reported by REVIGO when using SimRel).

| **GO Term** | **Name** |
| --- | --- |
| GO:0006511 | ubiquitin-dependent protein catabolic process |
| GO:0006513 | protein monoubiquitination |
| GO:0032020 | ISG15-protein conjugation |
| GO:0070979 | protein K11-linked ubiquitination |
| GO:0044314 | protein K27-linked ubiquitination |
| GO:0085020 | protein K6-linked ubiquitination |
| GO:0051865 | protein autoubiquitination |
| GO:1902042 | negative regulation of extrinsic apoptotic signaling pathway via death domain receptors |
| GO:0046330 | positive regulation of JNK cascade |
| GO:0043123 | positive regulation of canonical NF-kappaB signal transduction |
| GO:0010804 | negative regulation of tumor necrosis factor-mediated signaling pathway |
| GO:0042981 | regulation of apoptotic process |
| GO:0007249 | canonical NF-kappaB signal transduction |
| GO:0033209 | tumor necrosis factor-mediated signaling pathway |
| GO:0031571 | mitotic G1 DNA damage checkpoint signaling |
| GO:0042742 | defense response to bacterium |
| GO:0045087 | innate immune response |
| GO:0140374 | antiviral innate immune response |
| GO:0051092 | positive regulation of NF-kappaB transcription factor activity |
| GO:1902523 | positive regulation of protein K63-linked ubiquitination |
| GO:2000379 | positive regulation of reactive oxygen species metabolic process |

**Supplementary Table 2. Summary of the GO-BP terms that are enriched on gene data-driven pathway 11**, which is preferentially associated with MFAT-treated patients at timepoint T=6 months. We used REVIGO webserver to identify the most important GO terms (redundancy parameter =0.7). The selected GO terms are grouped together according to their semantic similarity (as reported by REVIGO when using SimRel).

| **GO Term** | **Name** |
| --- | --- |
| GO:0032897 | negative regulation of viral transcription |
| GO:0045892 | negative regulation of DNA-templated transcription |
| GO:0060271 | cilium assembly |

**Supplementary Table 3. Summary of the GO-BP terms that are enriched on gene data-driven pathway 23**, which is preferentially associated with MFAT-treated patients at timepoint T=6 months. We used REVIGO webserver to identify the most important GO terms (redundancy parameter =0.7). The selected GO terms are grouped together according to their semantic similarity (as reported by REVIGO when using SimRel).

| **GO Term** | **Name** |
| --- | --- |
| GO:0001556 | oocyte maturation |
| GO:0032467 | positive regulation of cytokinesis |
| GO:0010824 | regulation of centrosome duplication |
| GO:0051256 | mitotic spindle midzone assembly |
| GO:0000281 | mitotic cytokinesis |
| GO:0051591 | response to cAMP |
| GO:0090611 | ubiquitin-independent protein catabolic process via the multivesicular body sorting pathway |

**Supplementary Table 4. Summary of the GO-BP terms that are enriched on gene data-driven pathway 28**, which is preferentially associated with MFAT-treated patients at timepoint T=6 months. We used REVIGO webserver to identify the most important GO terms (redundancy parameter =0.7). The selected GO terms are grouped together according to their semantic similarity (as reported by REVIGO when using SimRel).

| **GO Term** | **Name** |
| --- | --- |
| GO:0006264 | mitochondrial DNA replication |
| GO:0000957 | mitochondrial RNA catabolic process |
| GO:1990481 | mRNA pseudouridine synthesis |
| GO:0006633 | fatty acid biosynthetic process |
| GO:0006783 | heme biosynthetic process |
| GO:0006552 | L-leucine catabolic process |
| GO:0032543 | mitochondrial translation |
| GO:0070681 | glutaminyl-tRNAGln biosynthesis via transamidation |
| GO:0070129 | regulation of mitochondrial translation |
| GO:1902775 | mitochondrial large ribosomal subunit assembly |
| GO:0032981 | mitochondrial respiratory chain complex I assembly |

**Supplementary Table 5. Summary of the GO-BP terms that are enriched on gene data-driven pathway 58**, which is preferentially associated with MFAT-treated patients at timepoint T=6 months. We used REVIGO webserver to identify the most important GO terms (redundancy parameter =0.7). The selected GO terms are grouped together according to their semantic similarity (as reported by REVIGO when using SimRel).

| **GO Term** | **Name** |
| --- | --- |
| GO:0032926 | negative regulation of activin receptor signaling pathway |
| GO:0045591 | positive regulation of regulatory T cell differentiation |
| GO:0002774 | Fc receptor mediated inhibitory signaling pathway |
| GO:0042129 | regulation of T cell proliferation |
| GO:1904948 | midbrain dopaminergic neuron differentiation |
| GO:0060070 | canonical Wnt signaling pathway |
| GO:0071294 | cellular response to zinc ion |
| GO:0071276 | cellular response to cadmium ion |
| GO:0140105 | interleukin-10-mediated signaling pathway |

**Supplementary Table 6. Summary of the GO-BP terms that are enriched on gene data-driven pathway 62**, which is preferentially associated with MFAT-treated patients at timepoint T=6 months. We used REVIGO webserver to identify the most important GO terms (redundancy parameter =0.7). The selected GO terms are grouped together according to their semantic similarity (as reported by REVIGO when using SimRel).

| **GO Term** | **Name** |
| --- | --- |
| GO:0016554 | cytidine to uridine editing |
| GO:0016556 | mRNA modification |
| GO:2000623 | negative regulation of nuclear-transcribed mRNA catabolic process, nonsense-mediated decay |

**Supplementary Table 7. Summary of the GO-BP terms that are enriched on gene data-driven pathway 67**, which is preferentially associated with MFAT-treated patients at timepoint T=6 months. We used REVIGO webserver to identify the most important GO terms (redundancy parameter =0.7). The selected GO terms are grouped together according to their semantic similarity (as reported by REVIGO when using SimRel).

| **GO Term** | **Name** |
| --- | --- |
| GO:0015760 | glucose-6-phosphate transport |
| GO:0015677 | copper ion import |
| GO:0035435 | phosphate ion transmembrane transport |
| GO:0042147 | retrograde transport, endosome to Golgi |
| GO:0030501 | positive regulation of bone mineralization |
| GO:0045648 | positive regulation of erythrocyte differentiation |
| GO:0045669 | positive regulation of osteoblast differentiation |
| GO:0036462 | TRAIL-activated apoptotic signaling pathway |
| GO:0007165 | signal transduction |
| GO:0007259 | cell surface receptor signaling pathway via JAK-STAT |
| GO:0032924 | activin receptor signaling pathway |
| GO:0035771 | interleukin-4-mediated signaling pathway |
| GO:0035772 | interleukin-13-mediated signaling pathway |
| GO:0038092 | nodal signaling pathway |
| GO:0038110 | interleukin-2-mediated signaling pathway |
| GO:0038113 | interleukin-9-mediated signaling pathway |
| GO:0060333 | type II interferon-mediated signaling pathway |
| GO:1990262 | anti-Mullerian hormone receptor signaling pathway |
| GO:0060021 | roof of mouth development |
| GO:0001649 | osteoblast differentiation |
| GO:0001837 | epithelial to mesenchymal transition |
| GO:0070723 | response to cholesterol |
| GO:0071773 | cellular response to BMP stimulus |
| GO:0099560 | synaptic membrane adhesion |
| GO:0106072 | negative regulation of adenylate cyclase-activating G protein-coupled receptor signaling pathway |
| GO:0032926 | negative regulation of activin receptor signaling pathway |
| GO:0060391 | positive regulation of SMAD protein signal transduction |

**Supplementary Table 8. Summary of the GO-BP terms that are enriched on gene data-driven pathway 70**, which is preferentially associated with MFAT-treated patients at timepoint T=6 months. We used REVIGO webserver to identify the most important GO terms (redundancy parameter =0.7). The selected GO terms are grouped together according to their semantic similarity (as reported by REVIGO when using SimRel).

| **GO Term** | **Name** |
| --- | --- |
| GO:0031669 | cellular response to nutrient levels |
| GO:0036295 | cellular response to increased oxygen levels |
| GO:0071230 | cellular response to amino acid stimulus |
| GO:0055062 | phosphate ion homeostasis |
| GO:0006879 | intracellular iron ion homeostasis |
| GO:0006885 | regulation of pH |
| GO:0007042 | lysosomal lumen acidification |
| GO:0030643 | intracellular phosphate ion homeostasis |
| GO:0061462 | protein localization to lysosome |
| GO:0006821 | chloride transport |
| GO:0006828 | manganese ion transport |
| GO:0006906 | vesicle fusion |
| GO:0008104 | intracellular protein localization |
| GO:0015818 | isoleucine transport |
| GO:0043308 | eosinophil degranulation |
| GO:0070254 | mucus secretion |
| GO:0070633 | transepithelial transport |
| GO:0070634 | transepithelial ammonium transport |
| GO:0072488 | ammonium transmembrane transport |
| GO:0072657 | protein localization to membrane |
| GO:0098708 | D-glucose import across plasma membrane |
| GO:0140361 | cyclic-GMP-AMP transmembrane import across plasma membrane |
| GO:0070072 | vacuolar proton-transporting V-type ATPase complex assembly |
| GO:0008361 | regulation of cell size |
| GO:0097254 | renal tubular secretion |
| GO:0150104 | transport across blood-brain barrier |
| GO:1902041 | regulation of extrinsic apoptotic signaling pathway via death domain receptors |
| GO:0032008 | positive regulation of TOR signaling |
| GO:2001200 | positive regulation of dendritic cell differentiation |
| GO:2000563 | positive regulation of CD4-positive, alpha-beta T cell proliferation |
| GO:1903076 | regulation of protein localization to plasma membrane |
| GO:1903593 | regulation of histamine secretion by mast cell |
| GO:1903595 | positive regulation of histamine secretion by mast cell |

**Supplementary Table 9. Summary of the GO-BP terms that are enriched on gene data-driven pathway 79**, which is preferentially associated with HA-treated patients at timepoint T=6 months. We used REVIGO webserver to identify the most important GO terms (redundancy parameter =0.7). The selected GO terms are grouped together according to their semantic similarity (as reported by REVIGO when using SimRel).

| **GO Term** | **Name** |
| --- | --- |
| GO:0032480 | negative regulation of type I interferon production |

**Supplementary Table 10. Summary of the GO-BP terms that are enriched on gene data-driven pathway 92**, which is preferentially associated with MFAT-treated patients at timepoint T=6 months. We used REVIGO webserver to identify the most important GO terms (redundancy parameter =0.7). The selected GO terms are grouped together according to their semantic similarity (as reported by REVIGO when using SimRel).

| **GO Term** | **Name** |
| --- | --- |
| GO:0031268 | pseudopodium organization |

**Supplementary Table 11. Summary of the GO-BP terms that are enriched on miRNA data-driven pathway 1**, which is preferentially associated with HA-treated patients at timepoint T=6 months. We used REVIGO webserver to identify the most important GO terms (redundancy parameter =0.7). The selected GO terms are grouped together according to their semantic similarity (as reported by REVIGO when using SimRel).

| **GO Term** | **Name** |
| --- | --- |
| GO:0006784 | heme A biosynthetic process |
| GO:0006004 | fucose metabolic process |
| GO:0006048 | UDP-N-acetylglucosamine biosynthetic process |
| GO:0006218 | uridine catabolic process |
| GO:0034626 | fatty acid elongation, polyunsaturated fatty acid |
| GO:0006534 | cysteine metabolic process |
| GO:0046293 | formaldehyde biosynthetic process |
| GO:0007076 | mitotic chromosome condensation |
| GO:0060155 | platelet dense granule organization |
| GO:1905691 | lipid droplet disassembly |
| GO:0010042 | response to manganese ion |
| GO:0000302 | response to reactive oxygen species |
| GO:0035456 | response to interferon-beta |
| GO:0010265 | SCF complex assembly |
| GO:0034380 | high-density lipoprotein particle assembly |
| GO:0017004 | cytochrome complex assembly |
| GO:0070925 | organelle assembly |
| GO:0014910 | regulation of smooth muscle cell migration |
| GO:0018021 | peptidyl-histidine methylation |
| GO:0030047 | actin modification |
| GO:0032886 | regulation of microtubule-based process |
| GO:0033625 | positive regulation of integrin activation |
| GO:0034241 | positive regulation of macrophage fusion |
| GO:1901098 | positive regulation of autophagosome maturation |
| GO:1903347 | negative regulation of bicellular tight junction assembly |
| GO:0033627 | cell adhesion mediated by integrin |
| GO:0035973 | aggrephagy |
| GO:0036151 | phosphatidylcholine acyl-chain remodeling |
| GO:0006629 | lipid metabolic process |
| GO:0042117 | monocyte activation |
| GO:0046602 | regulation of mitotic centrosome separation |
| GO:0048240 | sperm capacitation |
| GO:0001837 | epithelial to mesenchymal transition |
| GO:0002011 | morphogenesis of an epithelial sheet |
| GO:0009653 | anatomical structure morphogenesis |
| GO:0021599 | abducens nerve formation |
| GO:0035136 | forelimb morphogenesis |
| GO:0035261 | external genitalia morphogenesis |
| GO:0043586 | tongue development |
| GO:0048702 | embryonic neurocranium morphogenesis |
| GO:0048733 | sebaceous gland development |
| GO:0060218 | hematopoietic stem cell differentiation |
| GO:0060840 | artery development |
| GO:0061010 | gallbladder development |
| GO:0097187 | dentinogenesis |
| GO:2000793 | cell proliferation involved in heart valve development |
| GO:0051301 | cell division |
| GO:0051966 | regulation of synaptic transmission, glutamatergic |
| GO:0050855 | regulation of B cell receptor signaling pathway |
| GO:0070495 | negative regulation of thrombin-activated receptor signaling pathway |
| GO:2001045 | negative regulation of integrin-mediated signaling pathway |
| GO:0061755 | positive regulation of circulating fibrinogen levels |
| GO:0032848 | negative regulation of cellular pH reduction |
| GO:1905709 | negative regulation of membrane permeability |
| GO:0097753 | membrane bending |
| GO:0098586 | cellular response to virus |
| GO:0110025 | DNA strand resection involved in replication fork processing |
| GO:0006352 | DNA-templated transcription initiation |
| GO:0010792 | DNA double-strand break processing involved in repair via single-strand annealing |
| GO:0031118 | rRNA pseudouridine synthesis |
| GO:1901255 | nucleotide-excision repair involved in interstrand cross-link repair |
| GO:0055119 | relaxation of cardiac muscle |
| GO:0043116 | negative regulation of vascular permeability |
| GO:0050905 | neuromuscular process |
| GO:0060070 | canonical Wnt signaling pathway |
| GO:0007254 | JNK cascade |
| GO:0048011 | neurotrophin TRK receptor signaling pathway |
| GO:0070561 | vitamin D receptor signaling pathway |
| GO:0061744 | motor behavior |
| GO:0140706 | protein-containing complex localization to centriolar satellite |
| GO:0033344 | cholesterol efflux |
| GO:0035459 | vesicle cargo loading |
| GO:0036085 | GDP-fucose import into Golgi lumen |
| GO:0015810 | aspartate transmembrane transport |
| GO:0023061 | signal release |
| GO:0051640 | organelle localization |
| GO:0072383 | plus-end-directed vesicle transport along microtubule |
| GO:1902414 | protein localization to cell junction |
| GO:1902480 | protein localization to mitotic spindle |
| GO:1903093 | regulation of protein K48-linked deubiquitination |
| GO:1902004 | positive regulation of amyloid-beta formation |
| GO:1904093 | negative regulation of autophagic cell death |
| GO:0048147 | negative regulation of fibroblast proliferation |
| GO:1902255 | positive regulation of intrinsic apoptotic signaling pathway by p53 class mediator |
| GO:1903944 | negative regulation of hepatocyte apoptotic process |
| GO:1904775 | positive regulation of ubiquinone biosynthetic process |
| GO:1904973 | positive regulation of viral translation |
| GO:1900424 | regulation of defense response to bacterium |
| GO:0035491 | positive regulation of leukotriene production involved in inflammatory response |
| GO:1900036 | positive regulation of cellular response to heat |
| GO:1901509 | regulation of endothelial tube morphogenesis |
| GO:0010745 | negative regulation of macrophage derived foam cell differentiation |
| GO:0030500 | regulation of bone mineralization |
| GO:0045655 | regulation of monocyte differentiation |
| GO:0048638 | regulation of developmental growth |
| GO:0106089 | negative regulation of cell adhesion involved in sprouting angiogenesis |
| GO:1901331 | positive regulation of odontoblast differentiation |
| GO:1905381 | negative regulation of snRNA transcription by RNA polymerase II |
| GO:0010468 | regulation of gene expression |
| GO:0032909 | regulation of transforming growth factor beta2 production |
| GO:1901256 | regulation of macrophage colony-stimulating factor production |
| GO:1902894 | negative regulation of miRNA transcription |
| GO:1903427 | negative regulation of reactive oxygen species biosynthetic process |
| GO:2000097 | regulation of smooth muscle cell-matrix adhesion |
| GO:2000373 | positive regulation of DNA topoisomerase (ATP-hydrolyzing) activity |
| GO:2000299 | negative regulation of Rho-dependent protein serine/threonine kinase activity |

**Supplementary Table 12. Summary of the GO-BP terms that are enriched on miRNA data-driven pathway 12**, which is preferentially associated with MFAT-treated patients at timepoint T=6 months. We used REVIGO webserver to identify the most important GO terms (redundancy parameter =0.7). The selected GO terms are grouped together according to their semantic similarity (as reported by REVIGO when using SimRel).

| **Rank** | **Gene name** | **Link to OA** | **Evidence** |
| --- | --- | --- | --- |
| 1 | *OR52E6* | Indirect | PMID: 34746150 |
| 2 | *GOLGA8K* | Indirect | PMID: 38665758 |
| 3 | *LIMK2* | Direct | PMID: 25549868, PMID: 36849988 |
| 4 | *PPEF1* | Direct | PMID: 24229462 |
| 5 | *MOGAT1* | Indirect | PMID: 40405982 |
| 6 | *USP50* | Direct | EBI's GWAS catalog, GCST005813 |
| 7 | *OR10V1* | Indirect | PMID: 34746150 |
| 8 | *NEK11* | Indirect | PMID: 39693952 |
| 9 | *RGPD1* |  |  |
| 10 | *STK32C* | Direct | PMID: 36253145 |

**Supplementary Table 13. The top-10 genes that drive the preferential association of gene pathway 1** to the HA patients at timepoint T=6-months. For each gene, we report the evidence of its link to osteoarthritis (a direct link means that the gene is reported to be involved in OA in the literature, while an indirect link means that the function or the family of the gene is reported to be involved in OA in the literature).

| **Rank** | **Gene name** | **Link to OA** | **Evidence** |
| --- | --- | --- | --- |
| 1 | *SNORD88B* |  |  |
| 2 | *SNORD35A* | Direct | PMID: 32606371 |
| 3 | *SIRT7* | Direct | PMID: 33015765 |
| 4 | *SNORD60* |  |  |
| 5 | *SNORA10* |  |  |
| 6 | *SNORA11D* |  |  |
| 7 | *SNORD12C* |  |  |
| 8 | *SCARNA23* |  |  |
| 9 | *SNORA16A* |  |  |
| 10 | *SNORA5C* |  |  |

**Supplementary Table 14. The top-10 genes that drive the preferential association of gene pathway 62** to the MFAT patients at timepoint T=6-months. For each gene, we report the evidence of its link to osteoarthritis (a direct link means that the gene is reported to be involved in OA in the literature, while an indirect link means that the function or the family of the gene is reported to be involved in OA in the literature).

| **Rank** | **Gene name** | **Link to OA** | **Evidence** |
| --- | --- | --- | --- |
| 1 | *NDNF* |  |  |
| 2 | *FAM200B* |  |  |
| 3 | *IRX3* | Direct | PMID: 37433298 |
| 4 | *NDUFC1* | Direct | PMID: 41382139 |
| 5 | *SNORD6* |  |  |
| 6 | *SERTAD4-AS1* |  |  |
| 7 | *HS3ST3A1* |  |  |
| 8 | *SNORD2* |  |  |
| 9 | *SNORD75* |  |  |
| 10 | *SNHG6* |  |  |

**Supplementary Table 15. The top-10 genes that drive the preferential association of gene pathway 70** to the MFAT patients at timepoint T=6-months. For each gene, we report the evidence of its link to osteoarthritis (a direct link means that the gene is reported to be involved in OA in the literature, while an indirect link means that the function or the family of the gene is reported to be involved in OA in the literature).

| **Rank** | **Gene name** | **Link to OA** | **Evidence** |
| --- | --- | --- | --- |
| 1 | *RSPH10B* |  |  |
| 2 | *GCG* | Direct | PMID: 39995585 |
| 3 | *GID4* |  |  |
| 4 | *RANBP10* |  |  |
| 5 | *UBR3* | Indirect | PMID: 40917753 |
| 6 | *MMP13* | Direct | PMID: 33572320 |
| 7 | *MAEA* |  |  |
| 8 | *WDR26* |  |  |
| 9 | *ARMC8* |  |  |
| 10 | *RMND5A* | Indirect | PMID: 40917753 |

**Supplementary Table 16. The top-10 genes that drive the preferential association of gene pathway 79** to the HA patients at timepoint T=6-months. For each gene, we report the evidence of its link to osteoarthritis (a direct link means that the gene is reported to be involved in OA in the literature, while an indirect link means that the function or the family of the gene is reported to be involved in OA in the literature).

| **Rank** | **Gene name** | **Link to OA** | **Evidence** |
| --- | --- | --- | --- |
| 1 | *SKP2* | Direct | PMID: 38690715 |
| 2 | *SYT13* |  |  |
| 3 | *BTG1* |  |  |
| 4 | *NT5DC4* |  |  |
| 5 | *KLHDC1* |  |  |
| 6 | *CUL5* | Indirect | PMID: 34746150 |
| 7 | *COPS6* |  |  |
| 8 | *RCBTB1* |  |  |
| 9 | *SNORA67* |  |  |
| 10 | *FBXL20* |  |  |

**Supplementary Table 17. The top-10 genes that drive the preferential association of gene pathway 92** to the MFAT patients at timepoint T=6-months. For each gene, we report the evidence of its link to osteoarthritis (a direct link means that the gene is reported to be involved in OA in the literature, while an indirect link means that the function or the family of the gene is reported to be involved in OA in the literature).

| **Rank** | **miRNA name** | **Link to OA** | **Evidence** |
| --- | --- | --- | --- |
| 1 | HSA_MIR_7706 |  |  |
| 2 | HSA_MIR_337_5P | Direct | associated with OA in MirBase |
| 3 | HSA_LET_7D_3P | Indirect | associated with RA in MirBase |
| 4 | HSA_MIR_579_5P |  |  |
| 5 | HSA_MIR_574_3P | Indirect | associated with RA in MirBase |
| 6 | HSA_MIR_181A_3P |  |  |
| 7 | HSA_MIR_3605_3P |  |  |
| 8 | HSA_LET_7I_3P |  |  |
| 9 | HSA_MIR_1307_5P | Direct | associated with OA in MirBase |
| 10 | HSA_MIR_4746_5P |  |  |

**Supplementary Table 18. The top-10 miRNAs that drive the preferential association of miRNA pathway 1** to the HA patients at timepoint T=6-months. For each miRNA, we report the evidence of its link to osteoarthritis (a direct link means that the miRNA is reported to be involved in OA in MirBase, while an indirect link means that the miRNA is reported to be involved in a related disease, e.g, Rhumatoid Arthritis (RA) in MirBase).

| **Rank** | **miRNA name** | **Link to OA** | **Evidence** |
| --- | --- | --- | --- |
| 1 | HSA_MIR_6785_5P |  |  |
| 2 | HSA_MIR_4728_5P |  |  |
| 3 | HSA_MIR_6867_5P |  |  |
| 4 | HSA_MIR_1827 |  |  |
| 5 | HSA_MIR_7106_5P |  |  |
| 6 | HSA_MIR_6778_3P |  |  |
| 7 | HSA_MIR_3929 |  |  |
| 8 | HSA_MIR_6883_5P |  |  |
| 9 | HSA_MIR_6893_5P |  |  |
| 10 | HSA_MIR_4478 | Indirect | associated with RA in MirBase |

**Supplementary Table 19. The top-10 miRNAs that drive the preferential association of miRNA pathway 12** to the MFAT patients at timepoint T=6-months. For each miRNA, we report the evidence of its link to osteoarthritis (a direct link means that the miRNA is reported to be involved in OA in MirBase, while an indirect link means that the miRNA is reported to be involved in a related disease, e.g, Rhumatoid Arthritis (RA) in MirBase).
